# Combining multimorbidity clustering with limited demographic information enables high-precision outcome predictions

**DOI:** 10.1101/2024.05.28.24308024

**Authors:** Fabio S. Ferreira, Erwann Le Lannou, Benjamin Post, Shlomi Haar, Balasundaram Kadirvelu, Stephen J. Brett, A. Aldo Faisal

## Abstract

Multimorbidity, the coexistence of multiple health conditions in individuals, is prevalent and increasing worldwide, proving to be a growing challenge for patients and the healthcare systems. Furthermore, the prevalence of multimorbidity contributes to an increased risk of hospital admission or even death. In this study, we employ a principled approach that utilises longitudinal data routinely collected in electronic health records linked to half a million people from the UK biobank to generate digital comorbidity fingerprints (DCFs) using a topic modelling approach, Latent Dirichlet Allocation. These comorbidity fingerprints summarise a patient’s full secondary care clinical history, i.e. their comorbidities and past interventions. We identified 18 clinically relevant DCFs, which captured nuanced combinations of diseases and risk factors, e.g. grouping cardiovascular disorders with common risk factors but also novel groupings that are not obvious and differ in both their breadth and depth from existing observational disease associations. The DCFs, combined with demographic characteristics, performed on par or outperformed traditional models of all-cause mortality or hospital admission, showcasing the potential of data-driven strategies in healthcare forecasting. The comorbidity fingerprints together with age and number of hospital admissions were shown to be the most important factors in the predictions. Additionally, our DCF approach showed robust and consistent performance over time. Our findings underscore the promising role of interpretable data-driven approaches in healthcare forecasting, suggesting improved risk profiling for individual clinical decisions and targeted public health interventions, with consistent and robust performance over time.

**Author summary:** This study addresses the global challenge of multimorbidity, the presence of multiple health conditions in individuals, which is on the rise and poses a significant burden on patients and healthcare systems. Investigating its impact on the risk of hospitalization or mortality, we employ a sophisticated approach using longitudinal data from the UK Biobank to create digital comorbidity fingerprints (DCFs) through natural language processing methods. These DCFs, summarizing a patient’s complete clinical history, reveal 18 clinically relevant patterns, including unique combinations of diseases and risk factors. When combined with patient demographic and lifestyle data, the DCF approach performs similarly to traditional models in predicting all-cause mortality or hospitalization. Notably, the DCF approach demonstrates robust and consistent performance over time, highlighting its potential for enhancing healthcare forecasting. These findings emphasize the value of interpretable data-driven strategies in healthcare, offering improved risk profiling for individual clinical decisions and targeted public health interventions with enduring reliability.

## Introduction

Multimorbidity, the presence of two or more health conditions in an individual, is a widespread and escalating health challenge [1, 2]. Estimates of prevalence vary depending on the population examined and definitions used: in the UK ¿50% of people have at least two conditions by age 65, and ¿50% have more than three conditions by age 70 [3]. Additionally, the prevalence of multimorbidity is linked to poorer health outcomes, such as elevated mortality [4] and increased utilization of both inpatient and ambulatory healthcare services [5, 6]. The diverse nature of patients with multimorbidity results in a broad spectrum of condition combinations. Consequently, relying on overarching characterizations based solely on the count of conditions proves unhelpful in customising healthcare design. Recent studies suggest a shift from a simplistic disease-counting approach towards a more nuanced understanding of common co-occurring health conditions [2]. This transition aims to anticipate better the unique health needs and consequences of patients with specific combinations of conditions.

Hence, we consider a more principled approach to incorporating patient-specific information by systematically including comorbidities and their co-occurrence in a data-driven analysis to avoid human-induction bias, while carefully avoiding and signposting the potential for AI bias [7]. Our approach is enabled by large population datasets with linkage to registries such as death records and hospital admission and therefore represents a novel opportunity for automated clinical risk prediction model development on large cohorts. The use of such linked datasets has an established track record for the development and evaluation of clinical risk models, including those for cardiovascular disease, cancer, and mortality [8–11]. Here, we use the UK Biobank [12], which provides linked electronic health records data from the general population in the UK (it includes data from more than 500,000 people). A few studies have used data-driven approaches to identify multimorbidity clusters, but these included hospitalised patients only, mostly focusing on older age groups [13–16]. Additionally, the clusters’ interpretability in some cases depended on the preconditions selected by the researchers [16], and these studies do not validate the clusters on, for instance, predicting medical outcomes.

In this study, we utilise the longitudinal electronic health records from the UK Biobank to predict all-cause mortality and hospital admission. We demonstrate that the data already routinely collected and stored in EHRs can be rapidly leveraged to predict medical outcomes. Notably, we apply a topic modelling approach, Latent Dirichlet Allocation (LDA) [17, 18], on the entirety of the UK Biobank population to construct precise ‘digital comorbidity fingerprints’ (DCFs) from EHR data. We then test the validity of our approach by using the DCFs to train a Random Forrest Classifier to firstly predict 1-year, 2-year and 5-year all-cause mortality and secondly to predict 1-year and 2-year all-cause hospital admission. We demonstrate the face validity of such a two-step approach to model development by comparing the prediction performance of our DCFs, combined with previously documented demographic data, to a model based on hand-crafted feature selection previously proposed for all-cause mortality [9], a model based on hand-crafted feature selection for all-cause hospital admission (Post, B., et al., in preparation), and one model based entirely on demographic data. We further considered the interpretability of our data-driven DCFs and how they relate to the clinician-driven features and evaluated the robustness of our topic modelling approach by assessing its performance in a different timeframe and the stability of the identified DCFs. The approach proposed here is designed to provide a rapid, interpretable and accurate forecasting model for medical diseases based on past medical records.

## Methods

### Study population and data source

The UK Biobank is a large prospective population cohort of 502,505 participants aged 40-70 years at baseline, prepared to travel to 1 of 22 assessment centres in England, Scotland, and Wales [12]. All participants were recruited between 2006 and 2010, and all consented to have their health followed [12]. Baseline assessments include nurse-led interviews surrounding socio-demographics, lifestyle, physiological measurements, and medical history. Health outcomes and hospital admission data of the study participants were obtained from linkage to the Hospital Episode Statistics (HES) for England dataset, the Patient Episode Dataset for Wales (PEDW), the General Acute Inpatient and Day Case - Scottish Morbidity Record (SMR01), and the Mental Health Inpatient and Day Case - Scottish Morbidity Record (SMR04). Detailed information about the linkage procedure is available here. The latest linkage used in this study was completed in September 2020.

### Data preprocessing

Participants who were lost to follow-up (*n* = 1, 150) were removed from the final study population. Participants with missing ethnicity (*n* = 2, 740) were also dropped from the study as there is no reliable way to determine the right value for these participants. A total of 1,951 participants had already died before the start date of the study (i.e., 31 July 2010). The final study population included 496,664 individuals (Fig 1). Participants with missing smoking information were assumed to be non-smokers. Missing deprivation scores were imputed using the median value of the final study population.

**Fig 1.**
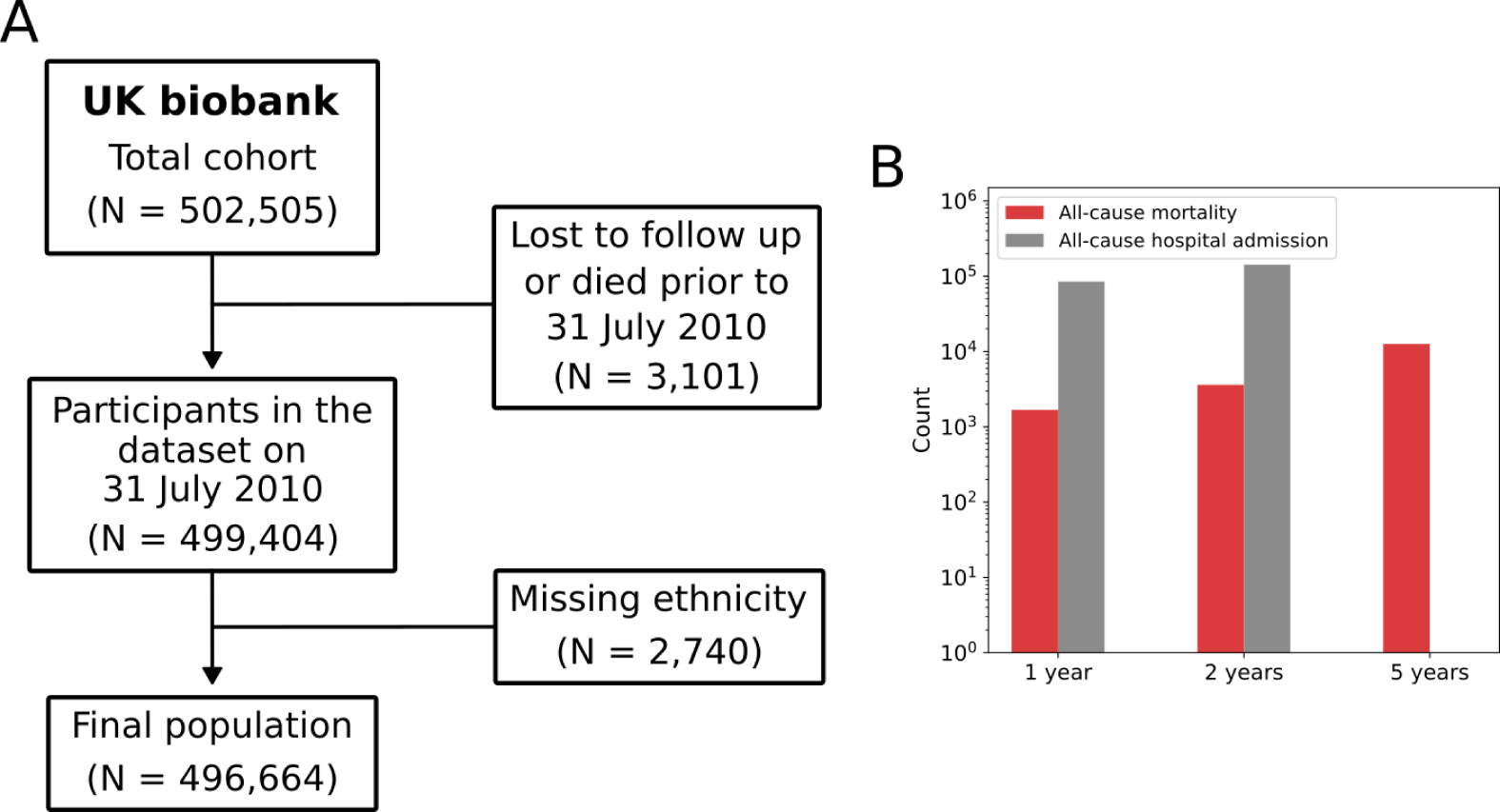
Cohort description. **(A)** Flowchart diagram of the study cohort. **(B)** Number of participants who died (in red) or had at least one hospital event (in grey) in one, two and five-year times (the 5-year outcome was only considered for the mortality outcome).

The hospital admission data were considered complete for our study population. For each participant, we considered a full longitudinal visit history to the hospital, including all admissions, emergency room attendances and outpatient appointments. For each recorded hospital episode, we extracted a list of diagnosis codes, using the ICD-10. This generated a document for each unique patient in the UK Biobank, representing their full hospital visit history as a series of ‘sentences’ (one per visit, so if a patient visits the hospital 10 times for a radiotherapy session it is represented 10 times in their record.), wherein each ‘word’ is a unique ICD-10 diagnosis code. Thus, this way of accounting naturally weighs conditions that require more frequent visits to the hospital.

### Baseline features

Demographic and lifestyle variables (i.e., age, sex, deprivation, ethnicity, and smoking status) were included as baseline features. Age was calculated for the index date chosen: 31 July 2010. Deprivation was measured using the Townsend Deprivation Index [19]. Ethnicity was grouped as white and non-white (thus collapsing the UK Biobank categories mixed, Asian, Black or Chinese), due to the otherwise small number of participants in each specific category. Smoking status was determined from data from the initial UK Biobank assessment and grouped into participants who currently smoke or have smoked (current or previous at baseline) and participants who, at baseline, were never smokers.

### Traditional model of hospital admission

The predictor features included for the all-cause hospital admission models were selected based on the features proposed by Post, B., et al. (in preparation) (summarised in S1 Table). The features related to the number of drug descriptions and GP events were not included here, as these were not available in our data. Each feature was constructed as a binary feature, indicating whether a given participant had in their past medical records the occurrence of at least one of the conditions for each feature. ICD-10 diagnosis codes were used to identify the underlying medical conditions.

### Digital comorbidity fingerprints (DCF): topic modelling

Topic modelling was originally developed as a tool to model collections of discrete data with particular applications in text modelling and natural language processing. Topic modelling can uncover latent ‘topics’ within a collection of documents, where a topic consists of a collection of words that frequently occur together. Here we used Latent Dirichlet Allocation (LDA), an unsupervised and interpretable method for topic modelling [17, 18]. LDA is a Bayesian probabilistic model that takes as input a corpus of documents and represents each document as a finite mixture of an underlying set of fixed topics, with each topic characterised by its distribution over words.

Here, LDA is used to represent a patient *p* as a mixture over the collection of *K* ‘topics’ (which we refer to, here, as a digital comorbidity fingerprint). Each topic *k* defines a multinomial distribution over a finite vocabulary of ICD-10 diagnosis codes and is assumed to have been drawn from a Dirichlet distribution, *β_k_∼* Dirichlet(*ν*). Thus, each code *c_p,n_* has a given probability *β_k_*in each topic *k*. Given the topics, LDA then generates for each patient *p* a distribution over topics *θ_p_ ∼* Dirichlet(*α*). In other words, each topic is determined as a distribution over closely related codes and each patient is represented as a distribution over multiple topics.

LDA was trained using a corpus of documents (i.e., sentences of ICD-10 diagnosis codes) for the entirety of the UK Biobank population that is not lost to follow-up and is still alive by the index date. This was chosen to generate more clinically robust and disease-agnostic topics (i.e., the topics formed are not specific to any prior comorbidity). Finally, we used a grid-search method to determine the optimal number *K* of topics, and the prior values of *θ* and *β* (*α* and *ν*, respectively). We optimise these hyperparameters using a normalised combined score of topic coherence [20] and perplexity, where we gave a higher weight to topic coherence as it has been shown that higher topic coherence improves topic interpretability [20, 21].

### Outcomes

We considered five health outcome measures: 1-year (i.e., the period between 31 July 2010 and 31 July 2011), 2-year (i.e., the period between 31 July 2010 and 31 July 2012) and 5-year all-cause mortality (i.e., the period between 31 July 2010 and 17 February 2016), and 1-year and 2-year all-cause hospital admission. All deaths were identified through linkage to national death registries, as explained here. The underlying cause(s) of death were determined by the International Classification of Diseases 10th Edition (ICD-10). A positive hospital admission outcome means that a given patient had had at least one hospital visit (e.g., inpatient appointment) during the chosen period. All outcomes are binary vectors, where one means the participant died/is admitted to the hospital and zero means the participant is alive/not admitted to the hospital.

### Random forest classifiers

We built three classifiers to predict the outcomes described in Outcomes, using three different sets of predictor features: demographics and lifestyle variables only (baseline model); demographics, lifestyle variables and derived LDA topics (DCF model); demographics, lifestyle variables and predictor features described in S1 Table (traditional model of hospital admission).

In this study, we used random forests (RF) as classifiers [22]. The RF algorithm involves fitting several decision tree classifiers on various sub-samples of the dataset and, subsequently, averaging these tree predictors. The random forest model’s hyperparameters control the learning function’s complexity and need to be tuned carefully. We tuned the hyperparameters using a grid search optimising for the AUROC (area under the receiver operating characteristic) curve on the validation sets. Since the data were extremely imbalanced, especially for the mortality-related outcomes (Fig. 1B), we balanced the training sets by randomly downsampling the negative class. The original imbalance of each outcome was kept the same for the validation and test sets, which were generated by randomly sampling from the remaining samples of each class. We report the mean and standard deviation of test AUROC values for all models, as a metric of the model’s performance.

The overview of the pipeline used for the DCF Model is illustrated in Fig. 2, which describes the learning of topics (i.e., digital comorbidity fingerprints) from the data and shows how we use the distribution of each patient over each of these topics as input to a classifier. We implemented all RF models using the *scikit-learn* library [23] in Python programming language.

**Fig 2.**
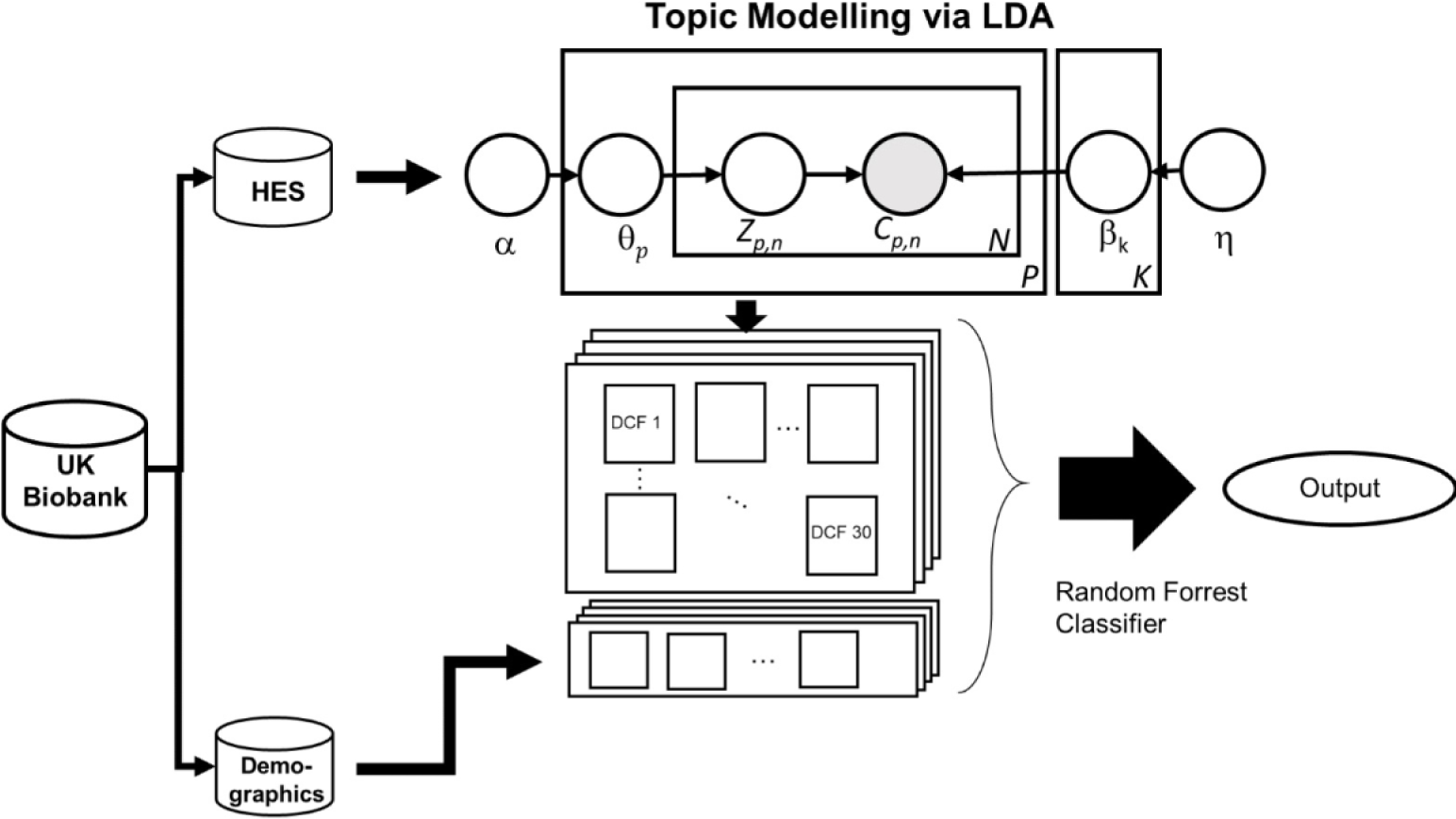
Illustration of the overall study design for the DCF Model. First, we separate the UK Biobank into HES data (all data for England, Scotland and Wales are considered here) and demographic data. We generate the digital comorbidity fingerprints from the ICD-10 code using LDA. We subsequently use the distribution of each patient over the DCFs as input to a random forest classifier to predict multiple outcomes (see Outcomes).

### Feature importance

To better understand the contribution of the features to each prediction, we used an impurity-based approach to rank the contribution of the different features in the prediction made by each model. This resulting feature importance score reflects the relative impact that each feature has on the model’s predictions. We used the RF algorithm for this analysis because it is a non-parametric algorithm that can recognise complex patterns, and automatically capture nonlinear and interaction effects without specifying these beforehand [24] while providing interpretable results.

### Statistical analysis

For each of the patient characteristics described in Traditional model of hospital admission, a logistic regression model was fitted with the 5-year all-cause mortality as the outcome to determine univariate (not-adjusted) Odds Ratio (OR). All patients’ characteristics were subsequently included in a single multivariate logistic regression to determine the adjusted OR. All ORs from the univariate and multivariate logistic regression models are reported with 95% confidence intervals (Table 1). In this study, the logistic regression model was not considered for its predictive power but rather for its ability to simply and accurately describe the available data.

**Table 1.** Descriptive characteristics of the UK Biobank cohort by 5-year all-cause mortality.

|  | **Died** (n = 12567) | **Alive** (n = 484097) | **OR** (95% CI) | **Fully adjusted OR** (95% CI) |
| --- | --- | --- | --- | --- |
| Age 50-59 | 3102 (24.68%) | 248813 (51.40%) | 1 | 1 |
| Age 60-69 | 7630 (60.71%) | 209778 (43.33%) | 2.92 (2.80 - 3.04) | 2.36 (2.26 - 2.47) |
| Age 70-79 | 1835 (14.60%) | 25506 (5.27%) | 5.77 (5.44 - 6.12) | 4.05 (3.81 - 4.32) |
| Atrial fibrillation | 784 (6.24%) | 7309 (1.51%) | 4.34 (4.02 - 4.68) | 1.44 (1.32 - 1.58) |
| Cancer | 2793 (22.22%) | 19963 (4.12%) | 6.64 (6.36 - 6.95) | 5.06 (4.82 - 5.30) |
| Chronic kidney disease | 214 (1.70%) | 823 (0.17%) | 10.2 (8.7 - 11.8) | 3.13 (2.63 - 3.73) |
| Chronic respiratory disease | 1480 (11.78%) | 21085 (4.36%) | 2.93 (2.77 - 3.10) | 1.60 (1.51 - 1.71) |
| Congestive cardiac failure | 221 (1.76%) | 621 (0.13%) | 13.9 (11.9 - 16.3) | 2.48 (2.06 - 2.98) |
| Connective tissue disease | 108 (0.86%) | 1313 (0.27%) | 3.19 (2.62 - 3.88) | 1.48 (1.19 - 1.84) |
| Depression | 400 (3.18%) | 5270 (1.09%) | 2.99 (2.69 - 3.31) | 1.50 (1.33 - 1.68) |
| Deprivation Q1 | 2645 (21.05%) | 121337 (25.06%) | 1 | 1 |
| Deprivation Q2 | 2769 (22.03%) | 121260 (25.05%) | 1.05 (0.99 - 1.11) | 1.03 (0.97 - 1.09) |
| Deprivation Q3 | 3113 (24.77%) | 121527 (25.10%) | 1.18 (1.12 - 1.24) | 1.16 (1.10 - 1.23) |
| Deprivation Q4 | 4040 (32.15%) | 119973 (24.78%) | 1.54 (1.47 - 1.62) | 1.44 (1.36 - 1.51) |
| Diabetes | 1252 (9.96%) | 12525 (2.59%) | 4.17 (3.92 - 4.43) | 1.74 (1.62 - 1.87) |
| Epilepsy | 242 (1.93%) | 2563 (0.53%) | 3.69 (3.23 - 4.21) | 1.95 (1.68 - 2.26) |
| Ethnicity (non-white) | 421 (3.35%) | 26478 (5.47%) | 0.60 (0.54 - 0.66) | 0.71 (0.64 - 0.78) |
| Falls | 776 (6.17%) | 13074 (2.70%) | 2.37 (2.20 - 2.55) | 1.44 (1.33 - 1.57) |
| Hemiplegia | 95 (0.76%) | 941 (0.19%) | 3.91 (3.16 - 4.83) | 1.46 (1.15 - 1.85) |
| Hypercholesterolemia | 1515 (12.06%) | 21221 (4.38%) | 2.99 (2.83 - 3.16) | 0.94 (0.88 - 1.01) |
| Hypertension | 3204 (25.50%) | 48019 (9.92%) | 3.11 (2.98 - 3.24) | 1.22 (1.16 - 1.29) |
| Ischaemic heart disease | 1496 (11.90%) | 17131 (3.54%) | 3.68 (3.48 - 3.90) | 1.24 (1.16 - 1.34) |
| Learning difficulties | 9 (0.07%) | 91 (0.02%) | 3.81 (1.92 - 7.56) | 1.49 (0.69 - 3.19) |
| Liver disease | 409 (3.25%) | 2487 (0.51%) | 6.51 (5.86 - 7.24) | 2.62 (2.32 - 2.96) |
| Mania or schizophrenia | 55 (0.44%) | 757 (0.16%) | 2.81 (2.13 - 3.69) | 1.81 (1.34 - 2.43) |
| Obesity | 312 (2.48%) | 4274 (0.88%) | 2.86 (2.54 - 3.21) | 1.09 (0.96 - 1.25) |
| Osteoarthritis | 1164 (9.26%) | 26044 (5.38%) | 1.80 (1.69 - 1.91) | 0.98 (0.92 - 1.05) |
| Osteoporosis | 287 (2.28%) | 3235 (0.67%) | 3.47 (3.07 - 3.92) | 1.32 (1.16 - 1.51) |
| Peptic ulcer disease | 32 (0.25%) | 297 (0.06%) | 4.16 (2.89 - 5.99) | 1.59 (1.06 - 2.38) |
| Peripheral vascular disease | 551 (4.38%) | 3171 (0.66%) | 6.95 (6.34 - 7.63) | 2.05 (1.84 - 2.28) |
| Rheumatoid arthritis or systemic lupus erythematosus | 226 (1.80%) | 3074 (0.63%) | 2.87 (2.50 - 3.28) | 1.65 (1.42 - 1.91) |
| Sex | 7559 (60.15%) | 218271 (45.09%) | 1.84 (1.77 - 1.91) | 1.65 (1.58 - 1.71) |
| Smoking status | 8833 (70.29%) | 285410 (58.96%) | 1.65 (1.58 - 1.71) | 1.33 (1.27 - 1.38) |
| Substance abuse | 741 (5.90%) | 6246 (1.29%) | 4.79 (4.43 - 5.18) | 2.11 (1.92 - 2.31) |
| Valvular heart disease | 398 (3.17%) | 3207 (0.66%) | 4.90 (4.41 - 5.45) | 1.56 (1.38 - 1.77) |
| Venous thromboembolic disease | 48 (0.38%) | 247 (0.05%) | 7.51 (5.51 - 10.24) | 2.55 (1.79 - 3.63) |

### Ethical approval and data availability

The UK Biobank study was approved by the North West Multi-Centre Research Ethics Committee (MREC) and the Community Health Index Advisory Group (CHIAG). All participants provided written informed consent before enrolment in the study. The UK Biobank protocol is available online [25]. Access to anonymised data for the UK Biobank cohort was granted by the UK Biobank Access Management Team (application number 21770). Ethical approval was granted by the National Research Ethics Committee (REC 16/NW/0274) for the overall UK Biobank cohort.

The UK Biobank cohort data supporting this study’s findings is available to researchers as approved by the Biobank Access Management Team. This work uses data provided by patients and collected by the NHS as part of their care and support.

### Code availability

The code will be released soon.

## Results

### Characteristics of the study population

A total of 496,664 participants from the UK biobank database were included in this study (Figure 1; cohort description in Table 1). Overall, the median age of the participants at baseline (on 31 July 2010) was 59.8 (range: 39.6 - 76.4) years, and 225,830 patients (45.5%) were male. The univariate and fully adjusted associations (calculated using logistic regression) between patient-level characteristics (Table 1) and odds of 5-year all-cause mortality (on 17 February 2016) are shown in Table 1. We used the patient characteristics described in S1 Table to characterise the impact of pre-existing medical conditions on the study population. The most common comorbidity in the deceased population was hypertension (25.5%) followed by cancer (22.2%) and hypercholesterolemia (12.1%). Increasing age showed a strong association with an increased likelihood of all-cause death, with participants aged over 70 years being over 4 times (fully adjusted odd ratio (OR): 4.05, 95% CI: 3.81 - 4.32) more likely to die compared to 50–59-year-olds (Table 1 and Fig. 3). Other significant characteristics that showed an increased likelihood of dying are a previous diagnosis of cancer (fully adjusted OR: 5.06, 95% CI: 4.82 - 5.30), chronic kidney disease (fully adjusted OR: 3.13, 95% CI:(2.63 - 3.73)) and liver disease (fully adjusted OR: 2.62, 95% CI:(2.32 - 3.73)).

**Fig 3.**
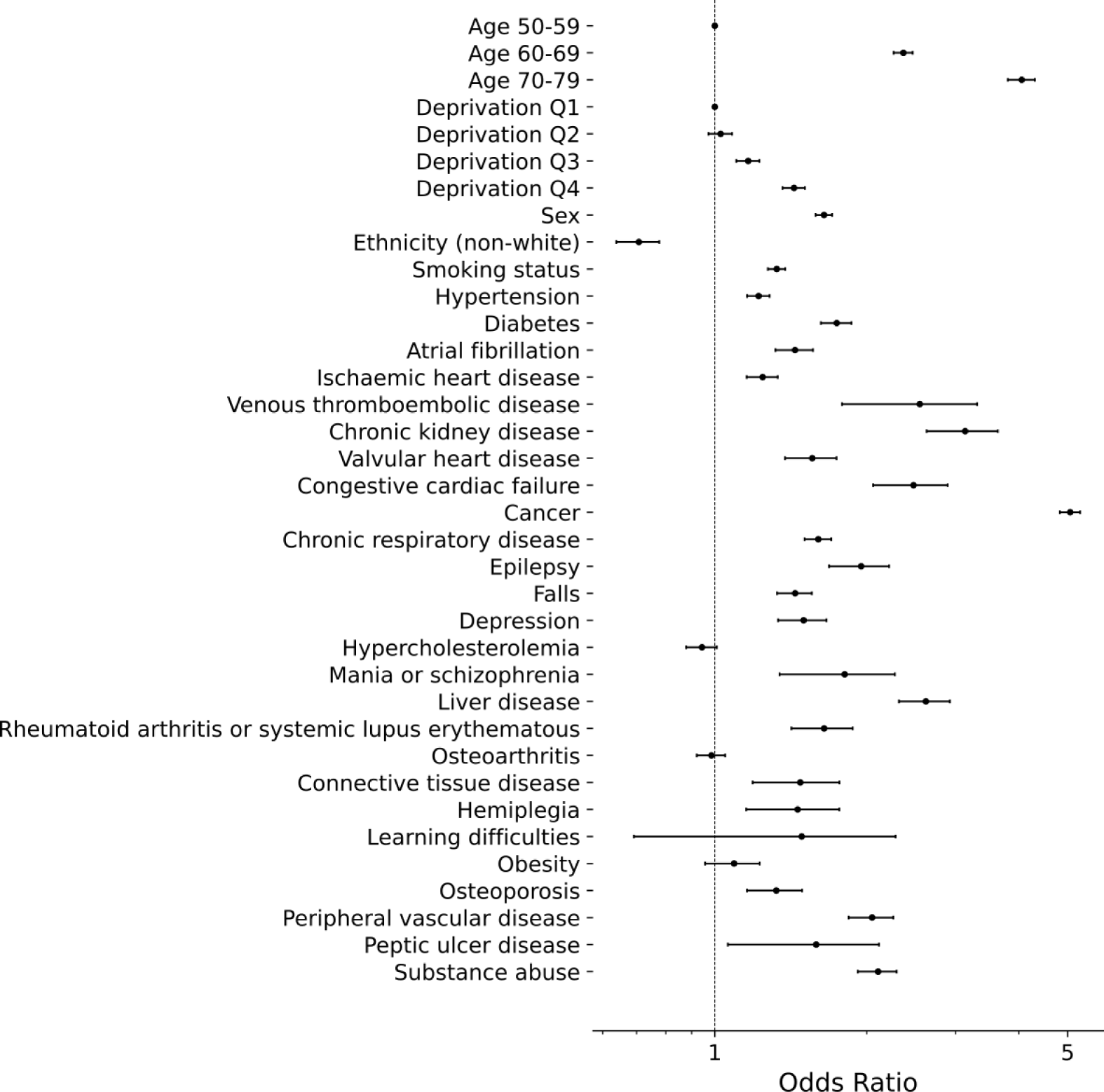
Estimated Odds Ratio for each patient characteristics. The adjusted Odds Ratio were estimated using a multivariate logistic regression. The patient characteristics are described in S1 Table

### Digital Comorbidity Fingerprinting (DCF)

Pre-existing conditions are coded as thousands of possible ICD-10 disease codes in patient records. Our methodology adapts unsupervised methods from natural language processing trained on the general population (i.e. the entire UK Biobank dataset) to cluster patients’ conditions using only disease codes from the patient’s past hospital visits. We used Latent Dirichlet Allocation (LDA) [17], a hierarchical Bayesian model that can naturally model the categorical nature and inherent imprecision of disease codes. This clustering distils thousands of disease codes, and billions of possible combinations into a set of topics, which we refer to here as Digital Comorbidity Fingerprints (DCFs). Each patient’s pre-existing conditions are thus summarised by a set of probabilities describing how strong a specific pre-existing comorbidity fingerprint is present in their record.

LDA was trained on ICD-10 codes of the 496,664 participants who were included from the UK Biobank cohort to generate the topic distributions over all patients (i.e., the DCFs) and the ICD-10 codes (i.e., the topic weights). The optimal number of topics (ranging from 2 to 50) and the best set of hyperparameters were optimised for maximising and minimising perplexity (see Digital comorbidity fingerprints (DCF): topic modelling). The resulting optimal number of topics was 18, i.e., all pre-existing conditions and their myriad combinations were summarised by a total of 18 DCFs.

We clinically reviewed disease codes grouped in each of the generated topics, i.e., the DCFs. We observe that our comorbidity fingerprinting does not group codes simply by the clinical features (e.g., renal disorders or diseases of the circulatory system) as can be seen in Fig. 4. Instead, LDA groups past clinical codes in a more functional way (the top-10 ICD-10 codes of each DCF are listed in S2 Table). For example, DCF 9 groups codes of cardiovascular disorders together (such as angina and chronic ischaemic heart disease), but also includes some of the most common risk factors for these diseases, such as hypercholesterolaemia, hypertension or family history of cardiac disorders.

**Fig 4.**
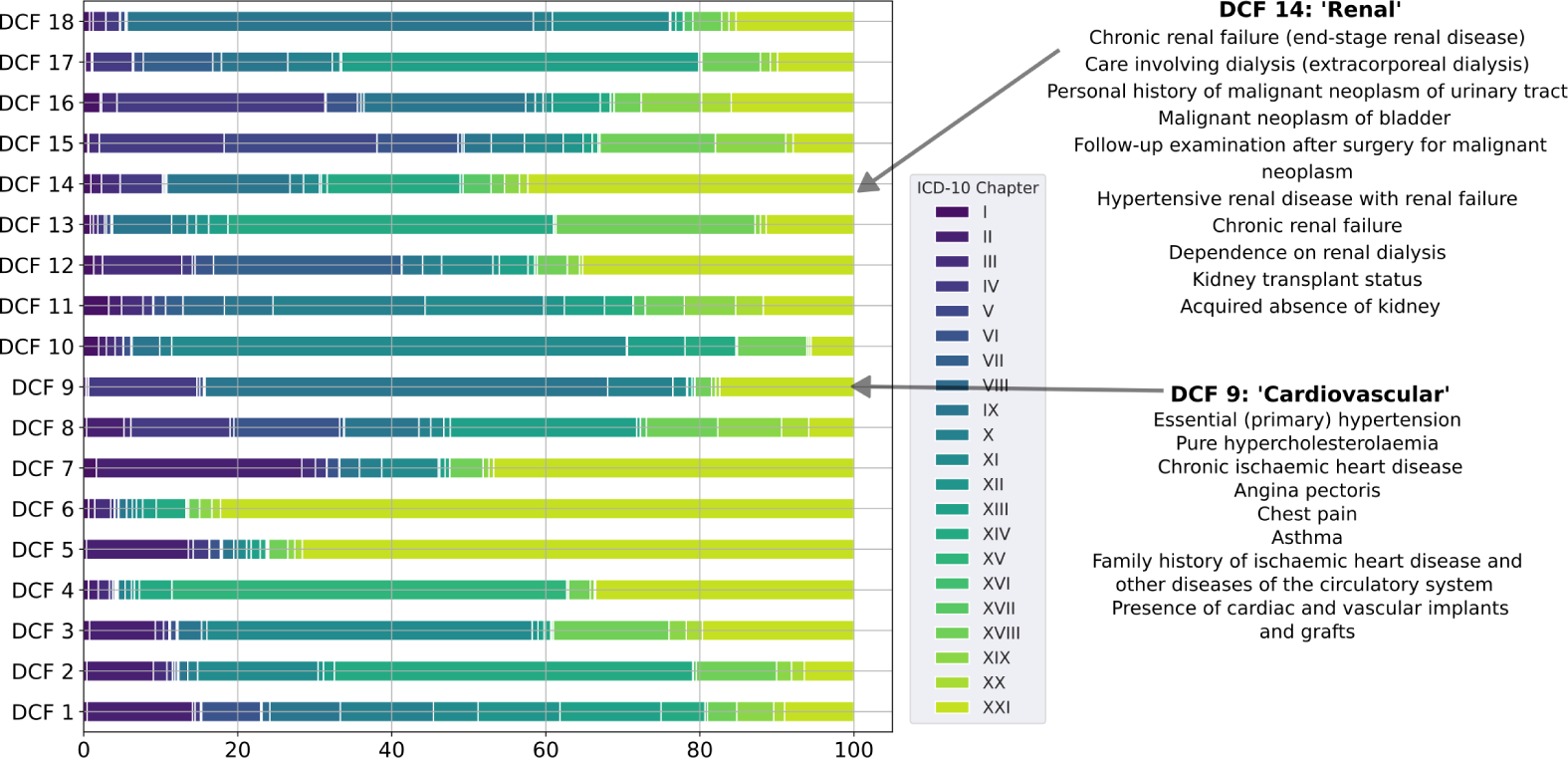
Distribution of ICD-10 code descriptions across the digital comorbidity fingerprints. An 18-topic LDA model was trained on the ICD-10 codes of the entire study population present in the dataset before 31 July 2010. On the left-hand side is the distribution of the codes over the ICD-10 ‘Chapters’ (see S1 Fig-S4 Fig) for all DCFs. On the right-hand side are shown the top ICD-10 code descriptions from DCF 9 and 14 (in rank order).

Additionally, DCF 16 groups diabetic disorders with occurrences of hypertension, hypercholesterolaemia and personal history of diseases of the circulatory system. Other fingerprints, such as DCF 5, DCF 6 and DCF 7, are mainly defined by neoplasms, chemotherapy sessions, radiotherapy sessions and surgical procedures (e.g., acquired absence of organs). The distribution of the proportion of words over the DCFs in the ICD-10 chapters is shown in S1 Fig-S4 Fig.

These results indicate that comorbidity fingerprinting can yield clinically relevant topics by grouping diseases that often occur simultaneously. The DCFs are further described in Fig. 5, which outlines the DCFs’ ability to encompass notions of gender (Fig. 5A), age (Fig. 5B) and body mass index (BMI) (Fig. 5C). For instance, DCF 4 which groups pregnancy, childbirth and the puerperium (traits that are normally associated with young adult women - see S3 FigC and S2 Table), was negatively correlated with age, and it was found to be more likely represented in women (Fig. 5A). DCF 9 (which groups cardiovascular disorders, hypercholesterolaemia and hypertension - see S2 Table) was positively correlated with age and BMI, and it is more likely related to men. DCF 16 (that groups diabetic disorders, hypercholesterolaemia and hypertension - see S2 Table) was also found positively correlated with age and BMI, and it has shown a greater likelihood of being related to men.

**Fig 5.**
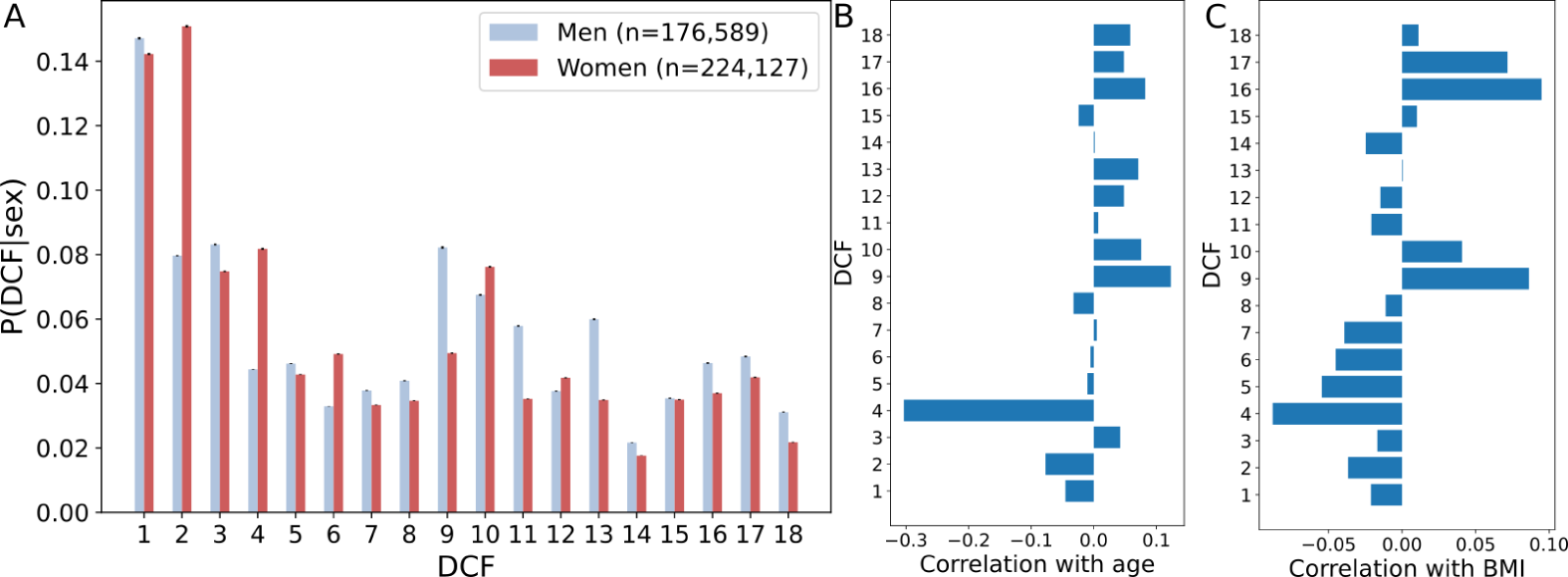
Summary of the Digital Comorbidity Fingerprints. **(A)** The average probability of belonging to each DCF by gender. **(B)** Pearson’s correlation between each DCF and age. **(C)** Pearson’s correlation between each DCF and body mass index (BMI).

### Prediction performance

We compared the prediction performance of the three proposed RF classifiers (see Random forest classifiers) in the all-cause mortality and hospital admission outcomes: (1) baseline model (using the set of features described in Baseline features); (2) traditional model (using the set of features described in Traditional model of hospital admission); (3) DCF model (using the DCFs obtained using LDA - see Digital comorbidity fingerprints (DCF): topic modelling). The prediction performance was evaluated in terms of the area under the receiver operating characteristic (AUROC) curve on the test sets. The DCF model performed better than the baseline and traditional models in all outcomes (index date: 31 July 2010) and performed on par with the model proposed by Weng et al. [9], which achieved an AUROC of 0.78 (95% CI: 0.78–0.79), for the 5-year all-cause mortality outcome (see Fig. 6 and S3 Table).

**Fig 6.**
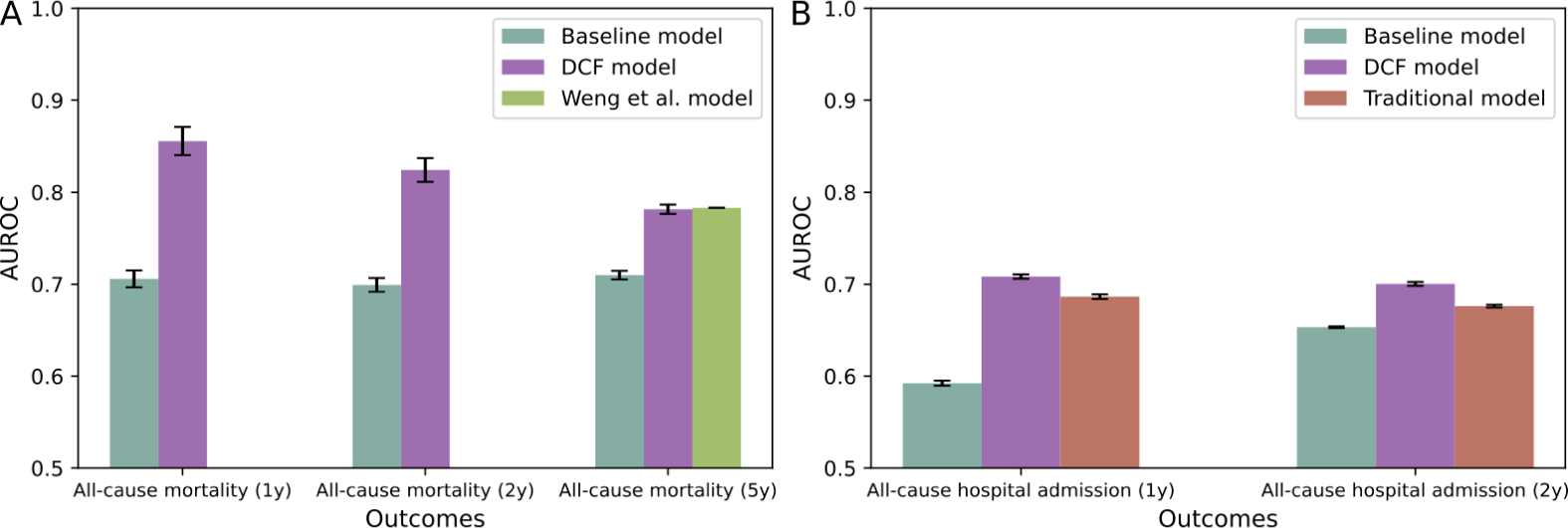
Performance of all prediction models for all mortality and hospital admission outcomes. **(A)** Mean AUROC for 1-year, 2-year and 5-year all-cause mortality on the test sets. **(B)** Mean AUROC for 1-year and 2-year all-cause hospital admission on the test sets. The error bars represent the standard deviation across five test sets.

### Model interpretability

In terms of the contributing factors to the all-cause mortality outcomes (Fig. 7 and S6 Fig), we observed that age and the DCF 4 were the two most important features. DCF 4 (which is related to young healthy women and pregnancy - see Fig. 5 and S2 Table) seems to be a protective factor, i.e., the participants who score high in it are less likely to die. DCF 2 (which also seems to be associated to women - Fig. 5 and S2 Table) also plays an important role, highlighting codes related to diseases of the genitourinary (S3 FigB) and digestive (S2 FigE) systems, and cancer (S1 FigB). DCF 1 and DCF 8 were identified as important factors and showed a more widespread distribution of the ICD-10 codes. The former groups a great variety of codes associated with diseases of the nervous (S1 FigF), respiratory (S2 FigD), circulatory (S2 FigC) and musculoskeletal (S3 FigA) systems, as well as skin diseases (S2 FigF) and cancer. The latter reflects preconditions related to diseases of the endocrine, nervous and musculoskeletal systems. Finally, fingerprints associated with neoplasms, chemotherapy sessions, radiotherapy sessions and surgical procedures (i.e., DCF 6 and 7), and renal diseases (DCF 14) played an important role as well.

**Fig 7.**
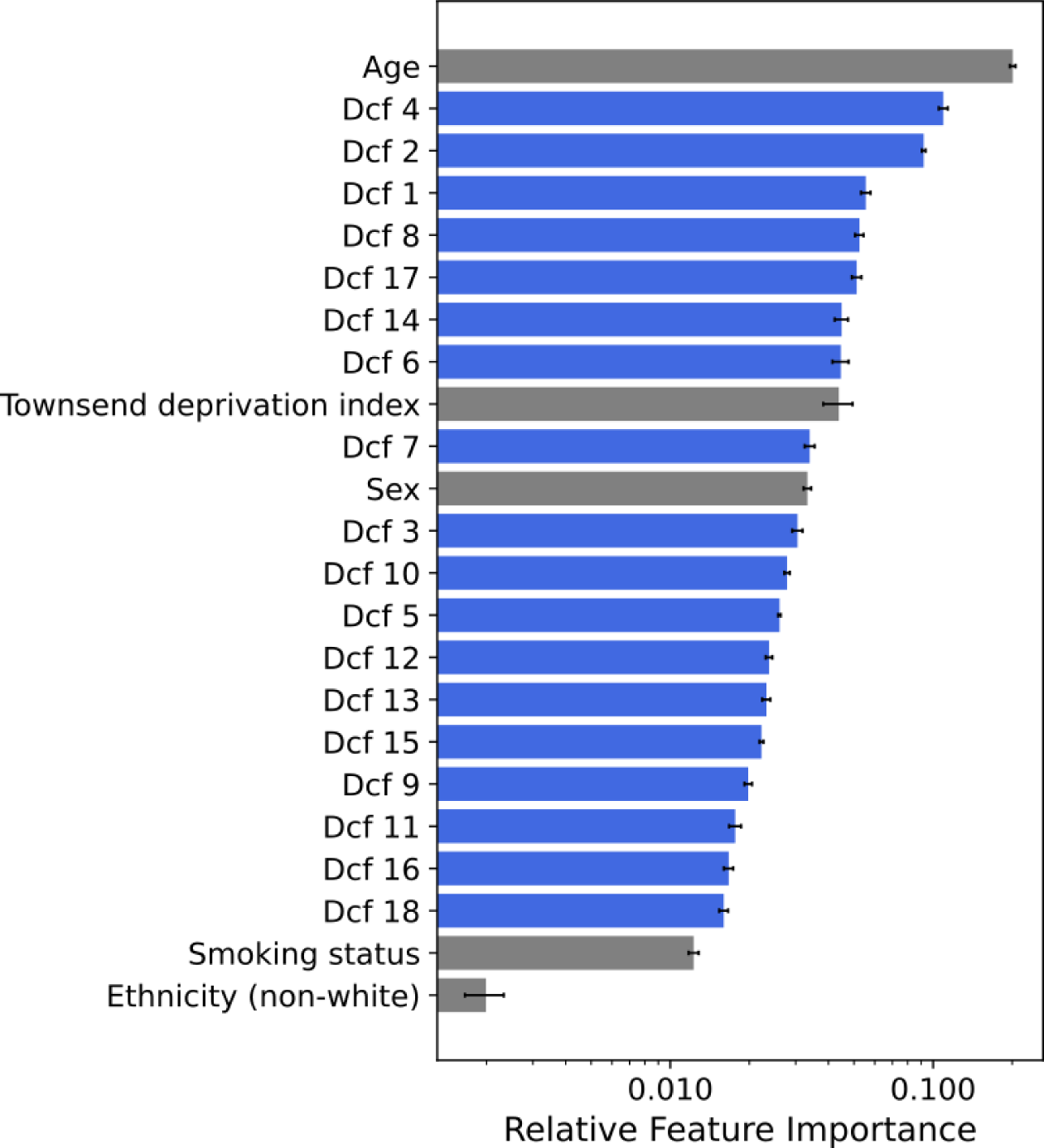
Feature importance of the DCF model for the 5-year all-cause mortality outcome. The grey bars show the relative importance of the demographics and lifestyle variables, whereas the blue bars show the relative importance of the digital comorbidity fingerprints (DCFs).

Regarding the features contributing the most to the 2-year all-cause hospital admission outcome (Fig. 8), we observed that whereas age and the prior number of hospital admissions are the most important features for the traditional model (Fig. 8A), this is not the case for the DCF model. DCF 4 (young healthy women and pregnancy), DCF 6 (breast cancer) and DCF 14 (renal disease) were the most relevant features for the DCF model’s predictions, followed by age, DCF 7 (multiple cancers) and the prior number of hospital admissions (Fig. 8B, see S2 Table and S1 Fig-S4 Fig for DCF interpretation). Deprivation also played an important role in the traditional model’s predictions (fourth most important feature), which was not observed in the DCF model (15th most important feature). The comorbidity-related features most important to the traditional model’s predictions comprised preconditions, such as hypertension, osteoarthritis, cancer, chronic respiratory disease, hypercholesterolemia, ischaemic heart disease and diabetes (Fig. 8A). The overall results were similar for the 1-year all-cause hospital admission with a slight reordering of the top most important features in both models (S5 FigA-B).

**Fig 8.**
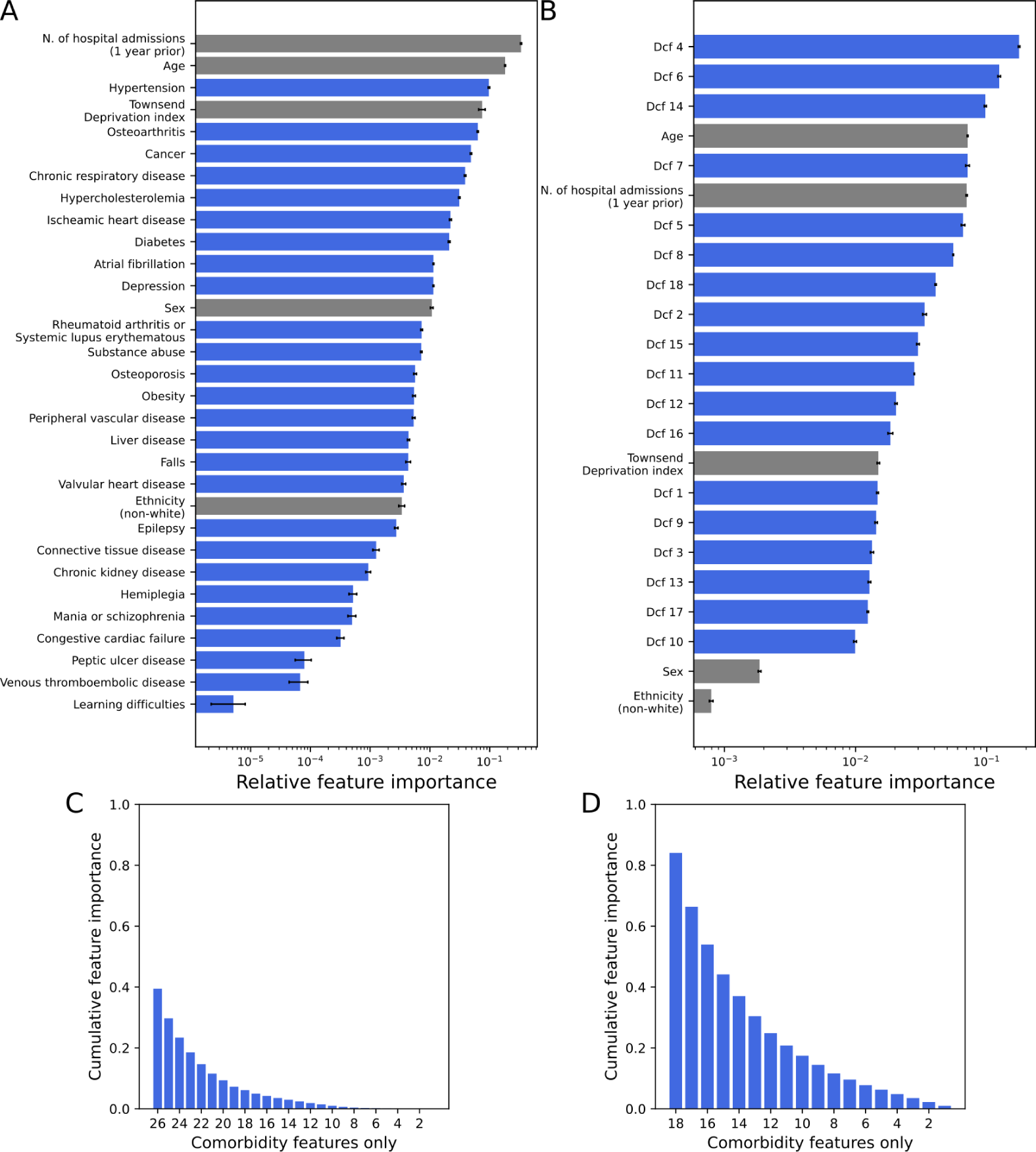
Feature importance of the prediction models for the 2-year all-cause hospital admission outcome. Relative feature importance of the **(A)** traditional model of hospital admission and **(B)** the DCF model. Cumulative feature importance of the **(C)** traditional model of hospital admission and **(D)** the DCF model. The grey bars show the relative importance of the demographics and lifestyle variables, whereas the blue bars show the relative importance of the comorbidity features.

In terms of total feature importance related to comorbidity features only, it can be seen that the DCFs played a significantly more important role (84% of cumulative feature importance) than the hand-crafted features (39% of cumulative feature importance) from the traditional model (Fig. 8C-D). A similar result was obtained for the 1-year all-cause hospital admission outcome: 81% and 39% of cumulative feature importance for the DCF and traditional model, respectively (S5 FigC-D).

### DCF model’s internal validation

To further evaluate the above-described methodology and its capacity to robustly inform decision-making at different time points, we used a similar approach, as described in Digital comorbidity fingerprints (DCF): topic modelling, to fit LDA to the ICD-codes of the entire population present in the dataset on 31 December 2014. The optimal number of DCFs identified was 22, which were then fed to an RF classifier to predict the same outcomes (described in Outcomes). The performance of all prediction models was similar to that shown for the index date of 31 July 2010, with the DCF model still performing the best for all outcomes (Fig. 9 and S4 Table). Additionally, we compared the 18 DCFs obtained on 31 July 2010 to the 22 DCFs obtained on 31 December 2014. S5 Table shows the maximum cosine similarity between each of the DCFs of the 2010 analysis and the most similar DCF of the 2014 analysis, where most comparisons achieved a cosine similarity greater than 0.5. For example, DCF 9 and 14 (Fig. 4) were found similar to DCF 10 and 20, respectively, in 2014, which grouped codes related to cardiovascular and renal disease as well (S7 Fig).

**Fig 9.**
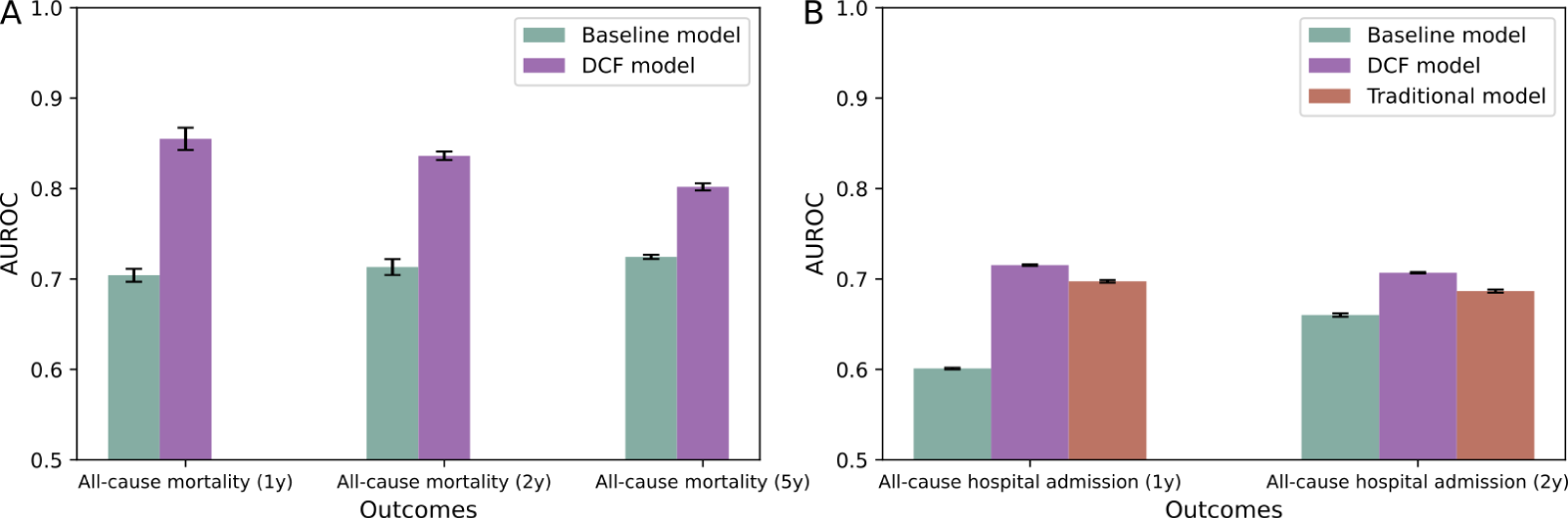
Performance of all prediction models for a different index date. **(A)** The test AUROCs for 1-year, 2-year and 5-year all-cause mortality. **(B)** The test AUROCs for 1-year and 2-year all-cause hospital admission. The values shown here correspond to the analysis with the index date of 31 December 2014.

## Discussion

Our findings demonstrate how taking a strictly data-driven approach using disease codes directly from electronic health records can be rapidly leveraged for predicting outcomes and studying disease patterns. The approach proposed here only required previously available data, based on rapidly collected patient diagnosis information and demographics, to predict who was likely to die or be admitted to a hospital in different time horizons. Even though our model does not rely on symptoms, laboratory values or images (at the time of diagnosis or during the illness), we can achieve comparable or higher predictive performance than traditional models of all-cause mortality and hospital admission.

To evaluate the utility of our topic model-based approach (i.e., the DCF model) to identify clinically interpretable dimensions, we compared it to previously proposed traditional risk models of all-cause mortality [9] and hospital admission (Post, B., et al., in preparation). Our method showed comparable discrimination performance to the all-cause mortality model and superior performance to the all-cause hospital admission model. The significance of our AI-based methodology is highlighted by the ability of our model to encompass the entire past medical history of the participants and not be restricted by potentially biased prior beliefs. In this way, our data-driven approach can avoid human induction bias in selecting the input features and, at the same time, may enable early stratification of key clinical risk groups thus permitting earlier intervention. Furthermore, we have shown here that our methodology is easily generalisable to multiple medical outcomes and robust to different timeframes, making it relevant for potential applicability in clinical practice.

Our study provides results regarding key findings in the underpinning structure of comorbidities and validates the usefulness of these associations with their predictability of mortality and hospital admission. The digital comorbidity fingerprints (DCFs) offer a rich and deep set of features, that can succinctly summarise a patient’s comorbidity profile using topic modelling. This approach allows for the use of a high-performance AI algorithm whilst remaining clinically interpretable and intuitive. Some obvious common causes of comorbidity clusters are well known (e.g. cardiovascular disease due to smoking or obesity [26]), advances in the science of multimorbidity are challenging because diseases may have linked or independent causes, combine heterogeneously, are treated differently, and thus affect care pathways and medication strategies. Structuring the myriad of combinations of comorbidities, such as through our comorbidity fingerprinting, underpins any measures to slow the accumulation of conditions and optimise their treatment. We have shown that AI methods can help in identifying multimorbidity clusters (i.e., the DCFs) without human-induction bias, while at the same time ensuring that findings remain clinically useful. Moreover, the DCFs showed great robustness to temporal shifts, as well as stable predictive performance over time which has been recently proposed as a more realistic and useful strategy to validate prediction models in clinical practice [27, 28].

We further demonstrate that the DCFs can effectively capture notions of well-known risk factors, such as hypertension, cancer or cardiovascular disease, as well as less straightforward factors like the protective factors grouped in DCF 4. When comparing the cumulative feature importance for the hospital admission outcomes, we observed that our comorbidity fingerprints played a more important role than the comorbidity features specified in the traditional model of hospital admission (Fig. 8), indicating that these data-driven comorbidity features can add value to predict multiple medical outcomes. However, we acknowledge that some DCFs may be more challenging to interpret and may not be as robust to temporal shifts, such as DCF 1 and DCF 8, which grouped a more diverse combination of preconditions.

There are a few limitations to our study. Firstly, our data set while vast may also reflect the inherent bias of the UK Biobank [29], which has been discussed in detail elsewhere [30, 31]; notably, the demographics reflect a “healthy volunteer” bias, with individuals being generally older, from more educated, less deprived socioeconomic backgrounds, and with significant under-representation of ethnic minorities compared to the UK population. In common with the UK biobank, our study is limited in that it does not fully represent the ethnic and socioeconomic diversity of the United Kingdom. Future work will be required before applying this approach in a more general context, specifically to assess better the risk in a younger and fully diverse population. Secondly, this study further relied on retrospective secondary care EHRs and the model is therefore currently blind to conditions entirely managed in primary care, such conditions include many less severe cases of diabetes, asthma and hypertension. At the time of the study, data from primary care were not available. Future work will be needed to incorporate data from General Practices into the development of the DCFs. Finally, we did not include the COVID-19 pandemic data, because, at the time of the study, we did not have access to data beyond 2020, which did not allow us to test our approach robustly beyond the pandemic. We were therefore unable to assess the considerable and irreversible change that the COVID-19 pandemic has caused in the national health service. Further research will be required to assess the robustness and performance of our approach in the case of novel diseases that have the potential to entirely and irreversibly disrupt the national health system, such as COVID-19.

In conclusion, we developed a novel data-driven approach to predict multiple medical outcomes in a community cohort using limited community EHR data. We have demonstrated the feasibility of a clustering approach comprising multimorbidities and long-term conditions. This clustering approach allows us to describe patient pre-conditions as belonging to just 18 clusters (DCFs) that avoid human-induction bias, often involving historical or speciality-dependent labels. This data-derived description enables us to succinctly summarise billions of possible comorbidity combinations, which we demonstrate as being effective in leveraging vast EHR data sets for medical outcome prediction. Our approach, applied at a population level, can assist in describing a risk profile in that population and is potentially useful in the provision of health care planning. Crucially, it may help in settings of rapid triaging such as emergency departments as patients or their carers would only have to provide basic demographic information and the DCF number that applies to them. This could transform accurate and rapid support of patients in health and care, as well as enable novel ways to reason at the public health level about patients. Our clustering approach opens up the question of how genetic, environmental and other contextual information may interplay to lead to these multimorbidity clusters.

## Supporting information

**S1 Fig.**
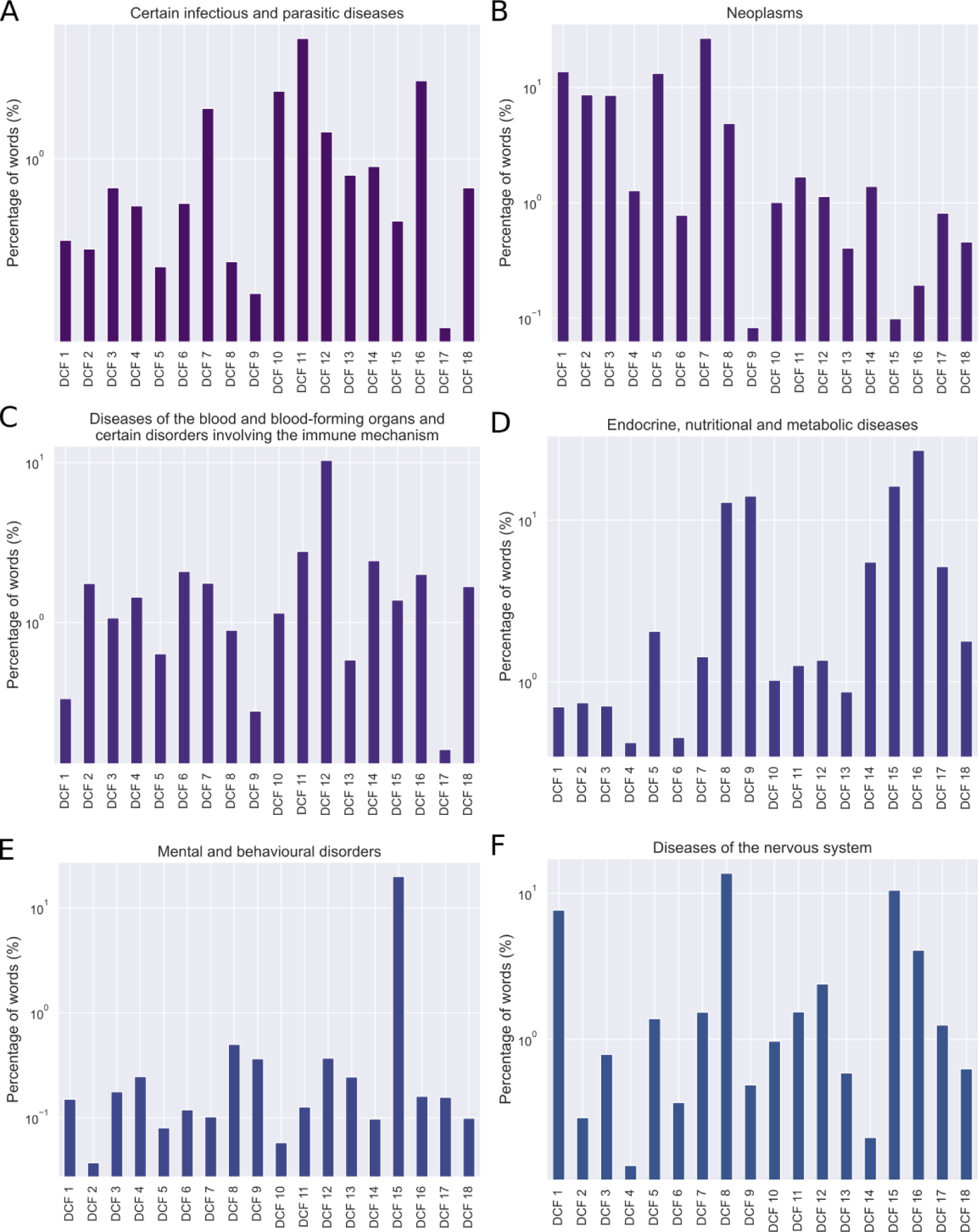
Distribution of the percentage of ICD-10 codes grouped in each DCF. Distribution of the percentage of codes from ICD-chapter **(A)** I, **(B)** II, **(C)** III, **(D)** IV, **(E)** V and **(F)** VI grouped in the DCFs obtained with data before 31 July 2010.

**S2 Fig.**
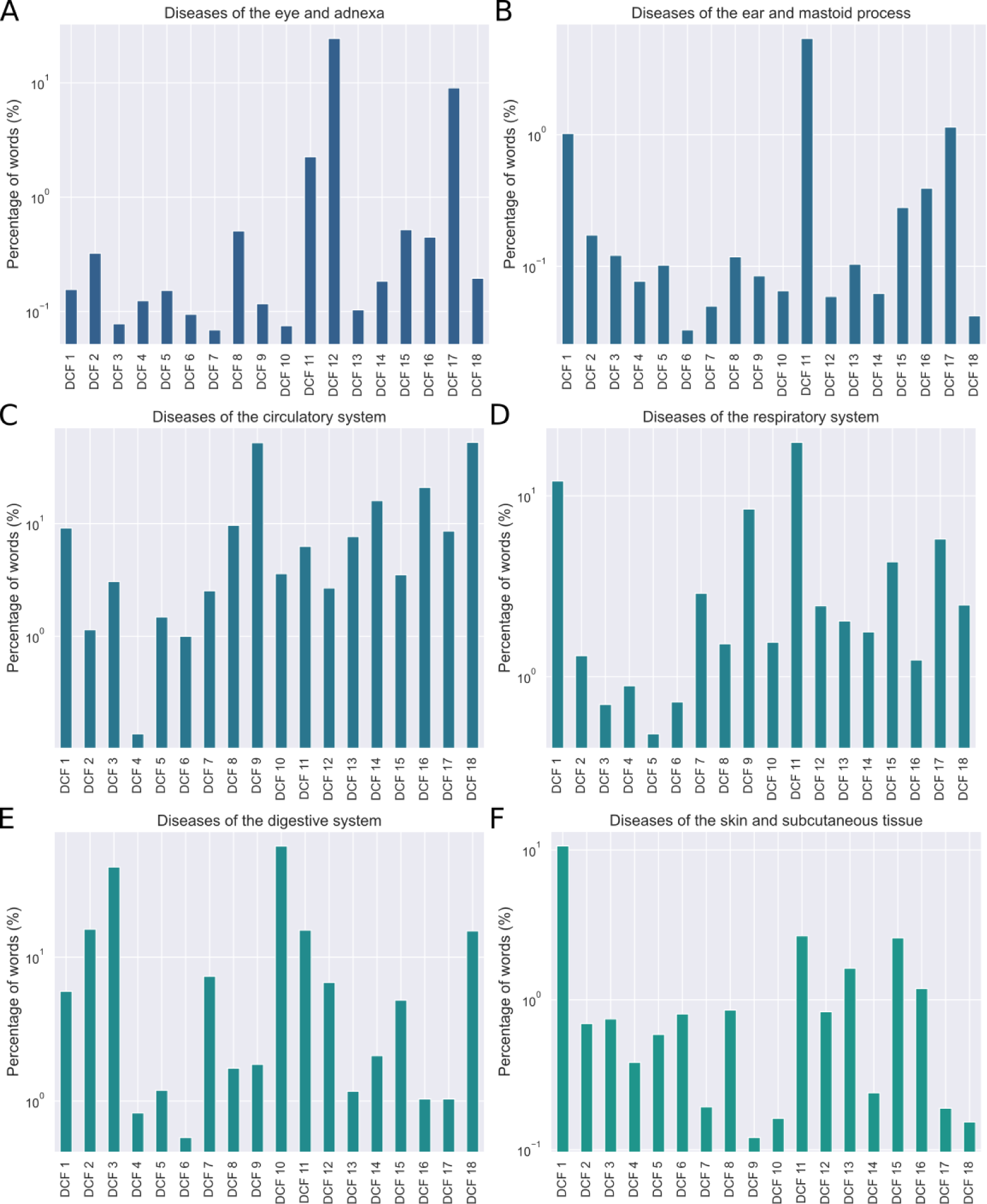
Distribution of the percentage of ICD-10 codes grouped in each DCF. Distribution of the percentage of codes from ICD-chapter **(A)** VII, **(B)** VIII, **(C)** IX, **(D)** X, **(E)** XI and **(F)** XII grouped in the DCFs obtained with data before 31 July 2010.

**S3 Fig.**
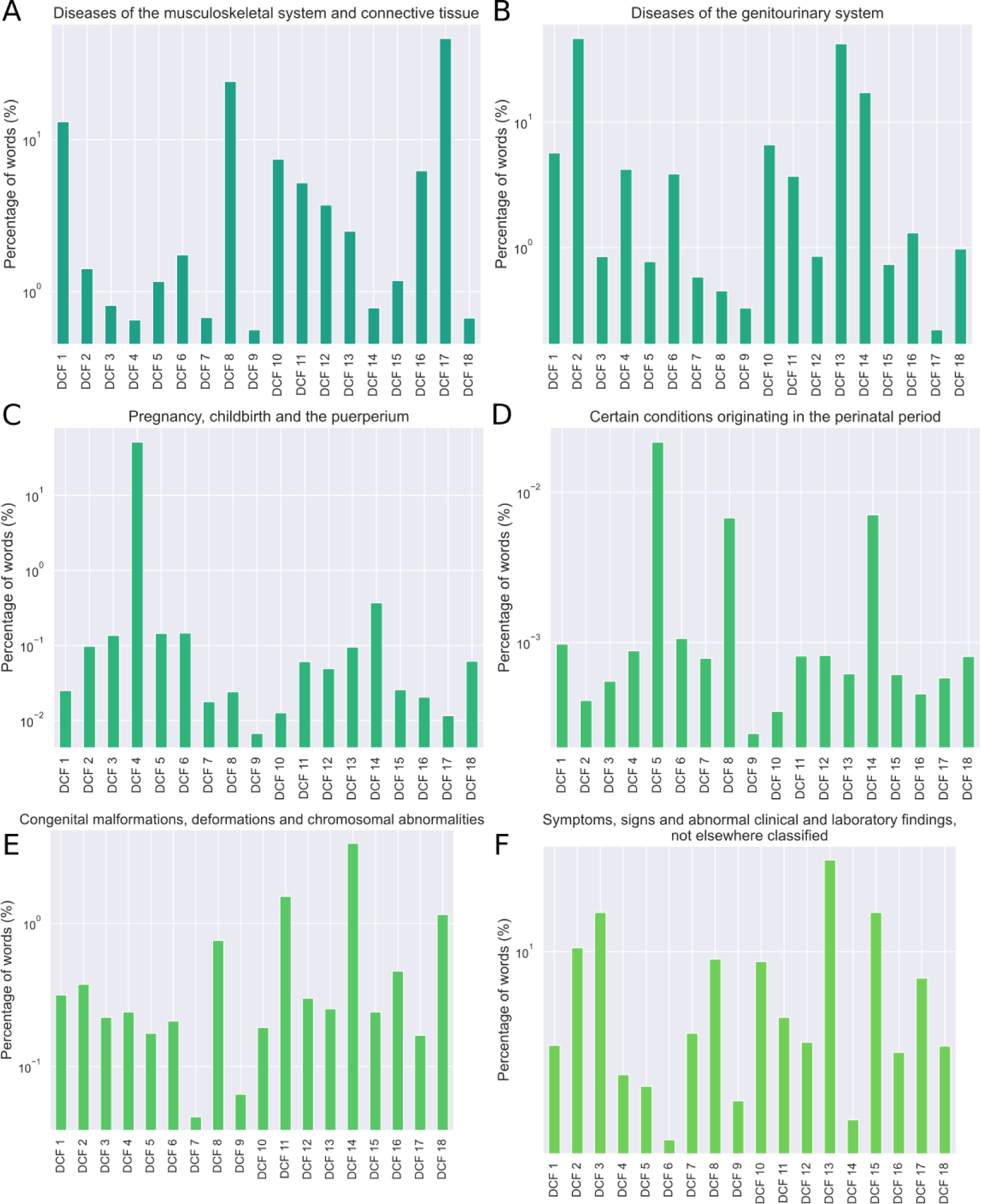
Distribution of the percentage of ICD-10 codes grouped in each DCF. Distribution of the percentage of codes from ICD-chapter **(A)** XIII, **(B)** XIV, **(C)** XV, **(D)** XVI, **(E)** XVII and **(F)** XVIII grouped in the DCFs obtained with data before 31 July 2010.

**S4 Fig.**
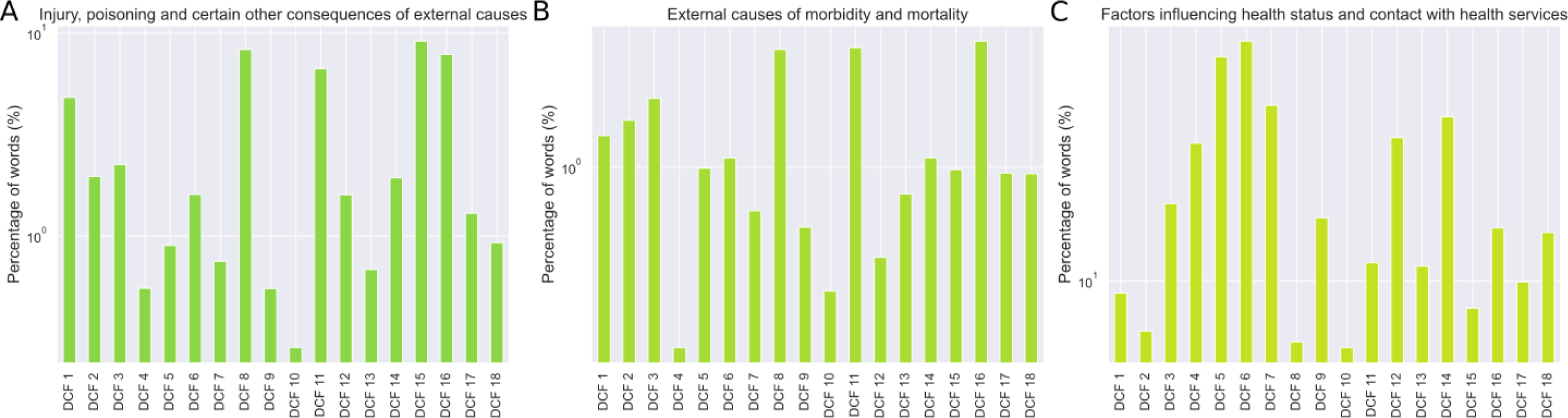
Distribution of the percentage of ICD-10 codes grouped in each DCF. Distribution of the percentage of codes from ICD-chapter **(A)** XIX, **(B)** XX, **(C)** XXI grouped in the DCFs obtained with data before 31 July 2010.

**S5 Fig.**
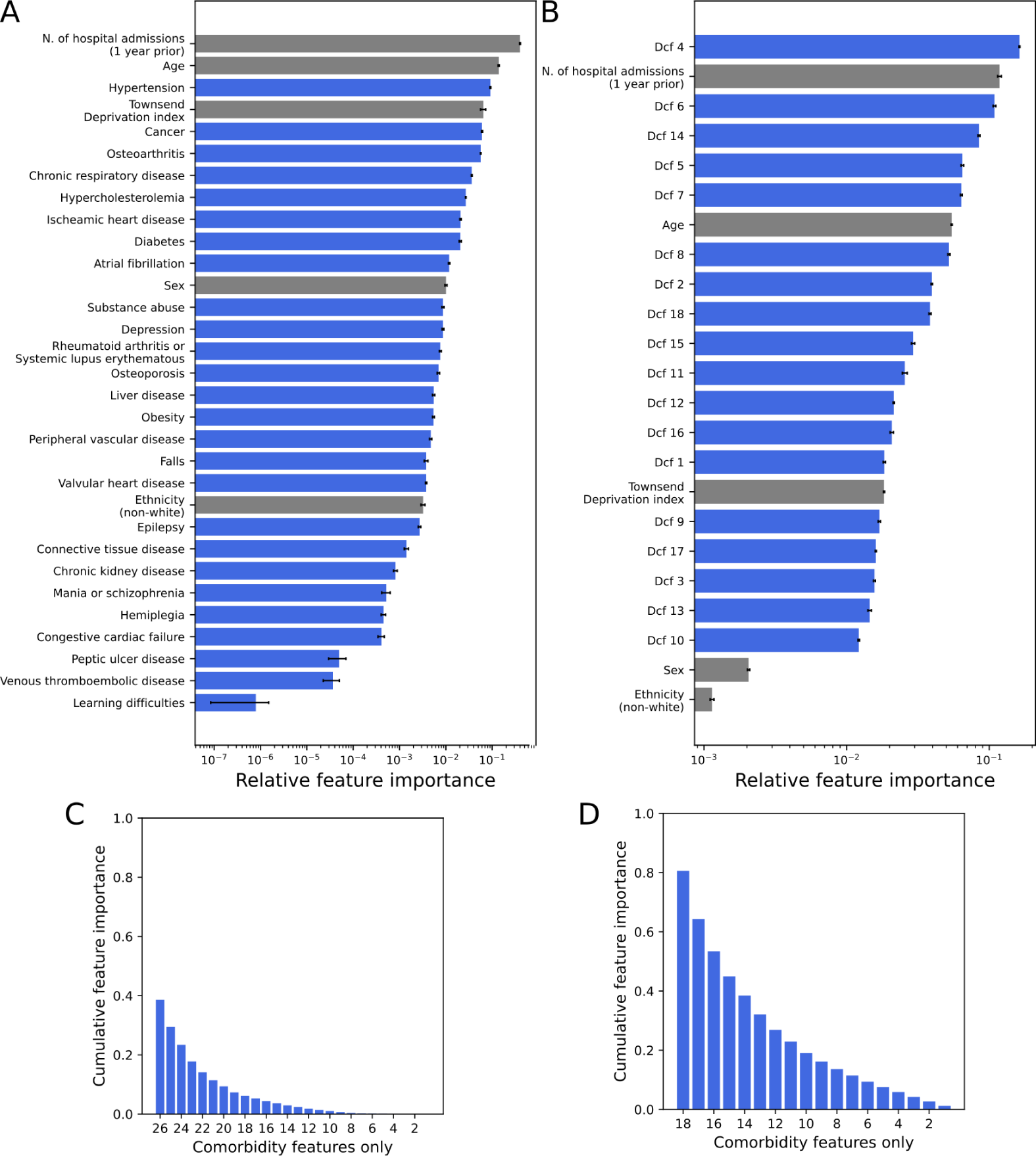
Feature importance of the prediction models for the 1-year all-cause hospital admission outcome. Relative feature importance of the **(A)** traditional model of hospital admission and **(B)** the DCF model. Cumulative feature importance of the **(C)** traditional model of hospital admission and **(D)** the DCF model. The grey bars show the relative importance of the demographics and lifestyle variables, whereas the blue bars show the relative importance of the comorbidity features.

**S6 Fig.**
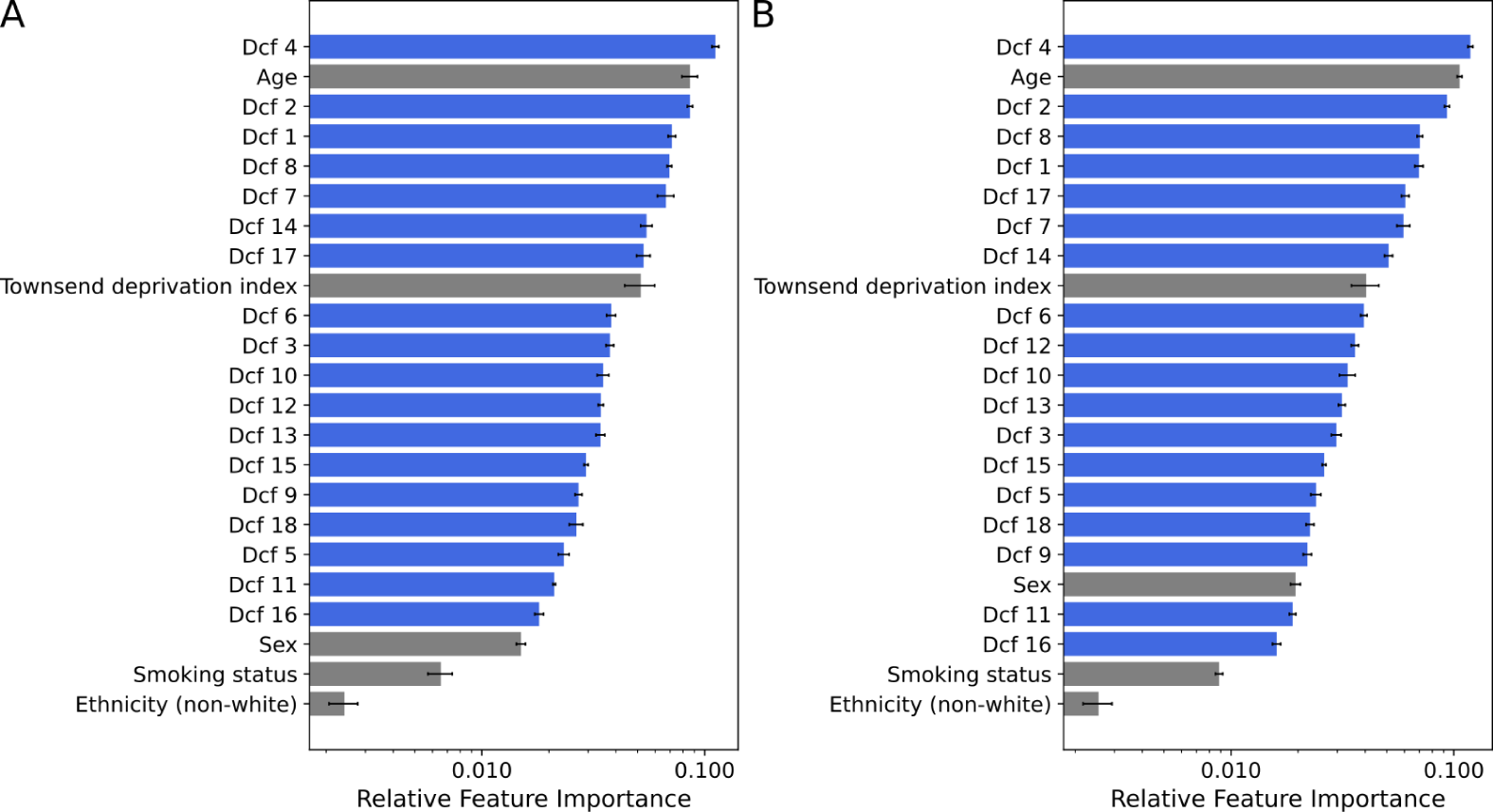
Feature importance of the DCF model for the (A) 1-year and (B) 2-year all-cause mortality outcomes. The grey bars show the relative importance of the demographics and lifestyle variables, whereas the blue bars show the relative importance of the DCFs.

**S7 Fig.**
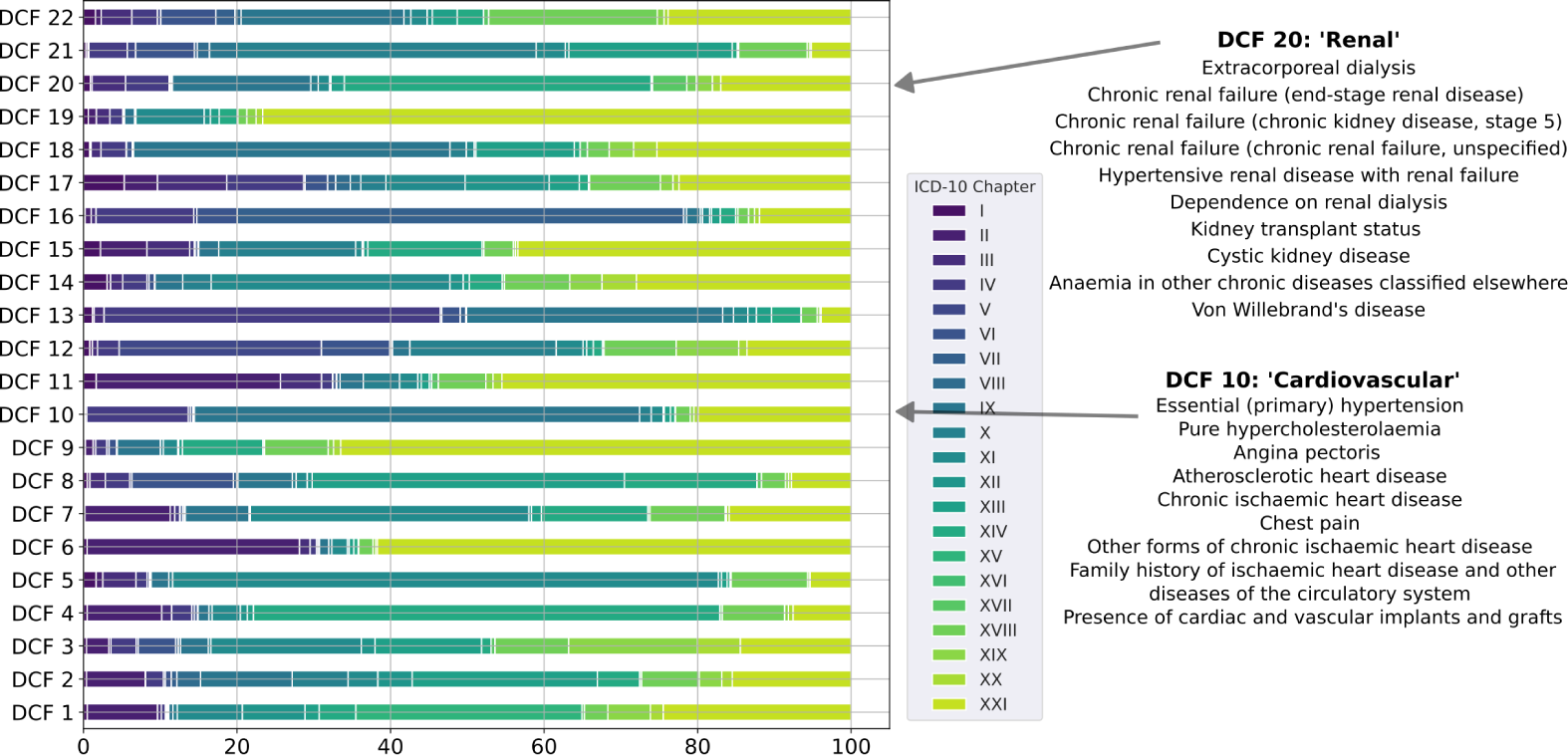
Distribution of ICD-10 code descriptions across the digital comorbidity fingerprints (31 December 2014). A 22-topic LDA model was trained on the ICD-10 codes of the entire study population present in the dataset before 31 December 2014. On the left-hand side is the distribution of the codes over the ICD-10 ‘Chapters’ (see S1 Fig-S4 Fig) for all DCFs. On the right-hand side are shown the top ICD-10 code descriptions from DCF 10 and 20 (in rank order).

**S1 Table.** Features included in the baseline and traditional models.

| Category | Description |
| --- | --- |
| Demographics | Age<br>Ethnicity<br>Sex<br>Townsend deprivation index |
| Lifestyle | Smoking status |
| Comorbidities | Atrial fibrillation<br>Cancer<br>Chronic kidney disease<br>Chronic respiratory disease<br>Congestive cardiac failure<br>Connective tissue disease<br>Depression<br>Diabetes<br>Epilepsy<br>Falls<br>Hemiplegia<br>Hypercholesterolemia<br>Hypertension<br>Ischaemic heart disease<br>Learning difficulties<br>Liver disease<br>Mania or schizophrenia<br>Obesity<br>Osteoarthritis<br>Osteoporosis<br>Peptic ulcer disease<br>Peripheral vascular disease<br>Rheumatoid arthritis or systemic lupus erythematosus<br>Substance abuse<br>Valvular heart disease<br>Venous thromboembolic disease |
| Total Hospital Admissions | Total hospital admissions in the 1 year before the index date |

**S2 Table.** Description of the top 10 ICD-10 codes in each DCF and its corresponding weights.

| DCF | ICD-10 code description | Weights |
| --- | --- | --- |
| 1 | Varicose veins of lower extremities without ulcer or inflammation | 0.051 |
|  | Mononeuropathies of upper limb (carpal tunnel syndrome) | 0.042 |
|  | Contraceptive management (sterilisation) | 0.030 |
|  | Acquired deformities of fingers and toes (hallux valgus) | 0.026 |
|  | Asthma | 0.025 |
|  | Follicular cysts of skin and subcutaneous tissue (trichilemmal cyst) | 0.021 |
|  | Persons encountering health services for specified procedures, not carried out | 0.018 |
|  | Essential (primary) hypertension | 0.018 |
|  | Other malignant neoplasms of skin (skin of other and unspecified parts of face) | 0.017 |
|  | Other disorders of nose and nasal sinuses (deviated nasal septum) | 0.017 |
| 2 | Excessive and frequent menstruation with a regular cycle | 0.046 |
|  | Leiomyoma of uterus | 0.045 |
|  | Menopausal and other perimenopausal disorders (postmenopausal bleeding) | 0.040 |
|  | Other diseases of anus and rectum (haemorrhage of anus and rectum) | 0.037 |
|  | Abdominal and pelvic pain (other and unspecified abdominal pain) | 0.036 |
|  | Polyp of female genital tract (polyp of corpus uteri) | 0.035 |
|  | Haemorrhoids | 0.028 |
|  | Non-infective gastro-enteritis and colitis | 0.020 |
|  | Noninflammatory disorders of ovary, Fallopian tube and broad ligament | 0.016 |
|  | Change in bowel habit | 0.016 |
| 3 | Unknown and unspecified causes of morbidity | 0.093 |
|  | Diverticular disease of large intestine without perforation or abscess | 0.074 |
|  | Personal history of diseases of the digestive system | 0.055 |
|  | Ulcerative colitis | 0.038 |
|  | Other diseases of intestine (polyp of colon) | 0.035 |
|  | Personal history of other neoplasms | 0.032 |
|  | Other diseases of anus and rectum (rectal polyp) | 0.025 |
|  | Other non-infective gastro-enteritis and colitis | 0.024 |
|  | Follow-up examination after surgery for other conditions | 0.024 |
|  | Diverticular disease of intestine | 0.023 |
| 4 | Outcome of delivery (single live birth) | 0.165 |
|  | Second-degree perineal laceration during delivery | 0.040 |
|  | First-degree perineal laceration during delivery | 0.026 |
|  | Dental caries | 0.020 |
|  | Single spontaneous delivery | 0.019 |
|  | Maternal care for other conditions predominantly related to pregnancy | 0.019 |
|  | Other abnormal products of conception (missed abortion) | 0.015 |
|  | Long labour | 0.015 |
|  | Maternal care due to uterine scar from previous surgery | 0.015 |
|  | Labour and delivery complicated by foetal heart rate anomaly | 0.015 |
| 5 | Radiotherapy session | 0.154 |
|  | Family history of malignant neoplasm of digestive organs | 0.113 |
|  | Malignant neoplasm of prostate | 0.074 |
|  | Personal history of malignant neoplasms of other organs and systems | 0.063 |
|  | Special screening examination for neoplasm of intestinal tract | 0.045 |
|  | Acquired absence of genital organ(s) | 0.027 |
|  | Persons encountering health services for other counselling and medical advice | 0.020 |
|  | Malignant neoplasm of rectosigmoid junction | 0.018 |
|  | Chemotherapy session for neoplasm | 0.016 |
|  | Malignant neoplasm of corpus uteri (endometrium) | 0.014 |
| 6 | Malignant neoplasm of breast | 0.246 |
|  | Chemotherapy session for neoplasm | 0.221 |
|  | Personal history of malignant neoplasm of breast | 0.098 |
|  | Secondary and unspecified malignant neoplasm of lymph nodes | 0.061 |
|  | Acquired absence of breast(s) | 0.035 |
|  | Malignant neoplasm of breast (upper-outer quadrant of breast) | 0.032 |
|  | Follow-up care involving plastic surgery of breast | 0.022 |
|  | Family history of malignant neoplasm of breast | 0.022 |
|  | Secondary malignant neoplasm of bone and bone marrow | 0.020 |
|  | Chronic lymphocytic leukaemia | 0.019 |
| 7 | Chemotherapy session for neoplasm | 0.179 |
|  | Personal history of malignant neoplasm of digestive organs | 0.060 |
|  | Malignant neoplasm of rectum | 0.046 |
|  | Malignant neoplasm of colon | 0.032 |
|  | Intra-abdominal lymph nodes | 0.031 |
|  | Secondary malignant neoplasm of liver | 0.028 |
|  | Malignant neoplasm of colon | 0.022 |
|  | Acquired absence of other parts of digestive tract | 0.022 |
|  | Diffuse non-Hodgkin's lymphoma (large cell) | 0.016 |
|  | Secondary malignant neoplasm of lung | 0.015 |
| 8 | Disorders of iron metabolism | 0.095 |
|  | Multiple sclerosis | 0.075 |
|  | Dorsalgia (low back pain) | 0.055 |
|  | Phlebitis and thrombophlebitis of other deep vessels of lower extremities | 0.039 |
|  | Polycythaemia vera | 0.037 |
|  | Unknown and unspecified causes of morbidity | 0.035 |
|  | Other chemotherapy | 0.021 |
|  | Dorsalgia (low back pain - site unspecified) | 0.021 |
|  | Dorsalgia (unspecified) | 0.019 |
|  | Pain in limb | 0.016 |
| 9 | Essential (primary) hypertension | 0.123 |
|  | Pure hypercholesterolaemia | 0.083 |
|  | Chronic ischaemic heart disease (atherosclerotic heart disease) | 0.077 |
|  | Angina pectoris | 0.070 |
|  | Chest pain | 0.061 |
|  | Asthma | 0.042 |
|  | Chronic ischaemic heart disease (other forms of chronic ischaemic heart disease) | 0.037 |
|  | Family history of ischaemic heart disease and other diseases of the circulatory system | 0.034 |
|  | Chronic ischaemic heart disease (unspecified) | 0.031 |
|  | Presence of cardiac and vascular implants and grafts | 0.029 |
| 10 | Diaphragmatic hernia | 0.100 |
|  | Gastritis and duodenitis | 0.044 |
|  | Gastro-oesophageal reflux disease with oesophagitis | 0.036 |
|  | Calculus of gallbladder without cholecystitis | 0.034 |
|  | Essential (primary) hypertension | 0.031 |
|  | Oesophagitis | 0.030 |
|  | Gastro-oesophageal reflux disease | 0.029 |
|  | Duodenitis | 0.025 |
|  | Dyspepsia | 0.025 |
|  | Abdominal and pelvic pain (pain localised to upper abdomen) | 0.025 |
| 11 | Inguinal hernia | 0.093 |
|  | Multiple myeloma and malignant plasma cell neoplasms | 0.052 |
|  | Cellulitis | 0.030 |
|  | Tobacco use | 0.028 |
|  | Asthma | 0.025 |
|  | Bronchiectasis | 0.018 |
|  | Transplanted organ and tissue status | 0.017 |
|  | Lobar pneumonia | 0.017 |
|  | Unspecified acute lower respiratory infection | 0.016 |
|  | Pulmonary embolism | 0.015 |
| 12 | Personal history of allergy to penicillin | 0.152 |
|  | Cataract | 0.101 |
|  | Non-Hodgkin's lymphoma | 0.039 |
|  | Iron deficiency anaemia | 0.036 |
|  | Personal history of allergy to analgesic agent | 0.027 |
|  | Coeliac disease | 0.022 |
|  | Essential (primary) hypertension | 0.019 |
|  | Personal history of allergy to other drugs, medicaments and biological substances | 0.018 |
|  | Personal history of allergy to other antibiotic agents | 0.016 |
|  | Irritable bowel syndrome | 0.016 |
| 13 | Unspecified haematuria | 0.080 |
|  | Hyperplasia of prostate | 0.063 |
|  | Essential (primary) hypertension | 0.052 |
|  | Other disorders of bladder | 0.032 |
|  | Other disorders of urinary system (urinary tract infection) | 0.031 |
|  | Rheumatoid arthritis | 0.030 |
|  | Retention of urine | 0.029 |
|  | Other chemotherapy | 0.027 |
|  | Calculus of kidney | 0.025 |
|  | Urethral stricture | 0.024 |
| 14 | Chronic renal failure (end-stage renal disease) | 0.231 |
|  | Care involving dialysis (extracorporeal dialysis) | 0.218 |
|  | Personal history of malignant neoplasm of urinary tract | 0.080 |
|  | Malignant neoplasm of bladder | 0.048 |
|  | Follow-up examination after surgery for malignant neoplasm | 0.043 |
|  | Hypertensive renal disease with renal failure | 0.043 |
|  | Chronic renal failure | 0.041 |
|  | Dependence on renal dialysis | 0.031 |
|  | Kidney transplant status | 0.018 |
|  | Acquired absence of kidney | 0.018 |
| 15 | Hypothyroidism | 0.109 |
|  | Depressive episode | 0.054 |
|  | Epilepsy | 0.045 |
|  | Syncope and collapse | 0.026 |
|  | Unknown and unspecified causes of morbidity | 0.023 |
|  | Mental and behavioural disorders due to use of alcohol (dependence syndrome) | 0.022 |
|  | Essential (primary) hypertension | 0.021 |
|  | Tobacco use | 0.020 |
|  | Psoriasis | 0.018 |
|  | Mental and behavioural disorders due to use of alcohol | 0.017 |
| 16 | Non-insulin-dependent diabetes mellitus | 0.139 |
|  | Essential (primary) hypertension | 0.111 |
|  | Personal history of diseases of the circulatory system | 0.060 |
|  | Insulin-dependent diabetes mellitus | 0.030 |
|  | Pure hypercholesterolaemia | 0.026 |
|  | Presence of orthopaedic joint implants | 0.022 |
|  | Coxarthrosis (arthrosis of hip) | 0.018 |
|  | Surgical operation with implant of artificial internal device | 0.016 |
|  | Coxarthrosis (other primary coxarthrosis) | 0.014 |
|  | Peripheral vascular disease | 0.010 |
| 17 | Gonarthrosis (arthrosis of knee) | 0.073 |
|  | Essential (primary) hypertension | 0.069 |
|  | Asthma | 0.042 |
|  | Gonarthrosis (other primary gonarthrosis) | 0.034 |
|  | Headache | 0.027 |
|  | Presence of orthopaedic joint implants | 0.027 |
|  | Diabetic retinopathy | 0.018 |
|  | Internal derangement of knee | 0.016 |
|  | Senile nuclear cataract | 0.014 |
|  | Shoulder lesions | 0.013 |
| 18 | Atrial fibrillation and flutter | 0.181 |
|  | Personal history of long-term (current) use of anticoagulants | 0.077 |
|  | Essential (primary) hypertension | 0.050 |
|  | Ulcer of oesophagus | 0.042 |
|  | Crohn's disease | 0.036 |
|  | Presence of cardiac pacemaker | 0.023 |
|  | Supraventricular tachycardia | 0.022 |
|  | Personal history of diseases of the circulatory system | 0.021 |
|  | Follicular non-Hodgkin's lymphoma | 0.021 |
|  | Nonrheumatic mitral valve disorders (valve insufficiency) | 0.017 |

**S3 Table.** Prediction performance for all mortality and hospital admission outcomes (31 July 2010). The values shown here refer to the test mean AUROC and its 95% confidence interval across five random splits of data. Bolded results indicate top-performing results per each outcome.

| Model | All-cause mortality |  |  | All-cause hospital admission |  |
| --- | --- | --- | --- | --- | --- |
|  | 1-year | 2-year | 5-year | 1-year | 2-year |
| Baseline | 0.70 (0.68-0.73) | 0.70 (0.68-0.72) | 0.71 (0.70-0.72) | 0.59 (0.58-0.60) | 0.65 (0.65-0.66) |
| Traditional | - | - | - | 0.69 (0.68-0.69) | 0.68 (0.67-0.68) |
| DCF | **0.86** (0.81-0.90) | **0.82** (0.79-0.86) | **0.78** (0.77-0.80) | **0.71** (0.70-0.71) | **0.70** (0.69-0.71) |

**S4 Table.** Prediction performance for all mortality and hospital admission outcomes (31 December 2014). The values shown here refer to the test mean AUROC and its 95% confidence interval across five random splits of data. Bolded results indicate top-performing results per each outcome.

| Model | All-cause mortality |  |  | All-cause hospital admission |  |
| --- | --- | --- | --- | --- | --- |
|  | 1-year | 2-year | 5-year | 1-year | 2-year |
| Baseline | 0.70 (0.68-0.72) | 0.71 (0.69-0.74) | 0.72 (0.72-0.73) | 0.60 (0.59-0.60) | 0.66 (0.66-0.67) |
| Traditional | - | - | - | 0.70 (0.69-0.70) | 0.69 (0.68-0.69) |
| DCF | **0.85** (0.82-0.89) | **0.84** (0.82-0.85) | **0.80** (0.79-0.81) | **0.72** (0.71-0.72) | **0.71** (0.71-0.71) |

**S5 Table.** Cosine similarity between the DCFs obtained before 31 July 2010 and before 31 December 2014. Values in bold represent cosine similarities greater than 0.60.

| DCF<br>(31 Jul 2010) | DCF<br>(31 Dec 2014) | Cosine similarity |
| --- | --- | --- |
| 1 | 2 | 0.387 |
| **2** | **4** | **0.795** |
| **3** | **7** | **0.649** |
| **4** | **1** | **0.828** |
| **5** | **9** | **0.670** |
| **6** | **19** | **0.979** |
| **7** | **6** | **0.967** |
| 8 | 16 | 0.297 |
| **9** | **10** | **0.962** |
| **10** | **5** | **0.891** |
| 11 | 17 | 0.385 |
| 12 | 2 | 0.447 |
| 13 | 7 | 0.395 |
| **14** | **20** | **0.901** |
| 15 | 12 | 0.501 |
| **16** | **13** | **0.897** |
| 17 | 2 | 0.568 |
| **18** | **18** | **0.857** |

## Data Availability

The UK Biobank cohort data supporting this study's findings is available to researchers as approved by the Biobank Access Management Team.

## Acknowledgments

We are grateful to the UK Biobank participants. This research has been conducted using the UK Biobank Resource under Application Number 21770. Infrastructure support for this research was provided by the Imperial NIHR Biomedical Research Centre. S.H. is supported by the Edmond and Lily Safra Fellowship Program and by the UK Dementia Research Institute Care Research & Technology Centre. AAF acknowledges generous funding from his UKRI Turing AI Fellowship (EP/V025449/1). The acknowledged parties and funders had no role in the design and outcome evaluation of the study.

## Notes

### Competing Interest Statement

The authors have declared no competing interest.

### Author Declarations

The UK Biobank study was approved by the North West Multi-Centre Research Ethics Committee (MREC) and the Community Health Index Advisory Group (CHIAG). All participants provided written informed consent before enrolment in the study. Access to anonymised data for the UK Biobank cohort was granted by the UK Biobank Access Management Team (application number 21770). Ethical approval was granted by the National Research Ethics Committee (REC 16/NW/0274) for the overall UK Biobank cohort.

